# Metformin on Time to Sustained Recovery in Adults with COVID-19: The ACTIV-6 Randomized Clinical Trial

**DOI:** 10.1101/2025.01.13.25320485

**Authors:** Carolyn T. Bramante, Thomas G. Stewart, David R. Boulware, Matthew W. McCarthy, Yue Gao, Russell L. Rothman, Ahmad Mourad, Florence Thicklin, Jonathan B. Cohen, Idania T. Garcia del Sol, Juan Ruiz-Unger, Nirav S. Shah, Manisha Mehta, Orlando Quintero Cardona, Jake Scott, Adit A. Ginde, Mario Castro, Dushyantha Jayaweera, Mark Sulkowski, Nina Gentile, Kathleen McTigue, G. Michael Felker, Sean Collins, Sarah E. Dunsmore, Stacey J. Adam, Christopher J. Lindsell, Adrian F. Hernandez, Susanna Naggie, the Accelerating COVID-19 Therapeutic Interventions and Vaccines (ACTIV)-6 Study Group and Investigators

## Abstract

**Importance:** The effect of metformin on reducing symptom duration among outpatient adults with coronavirus disease 2019 (COVID-19) has not been studied.

**Objective:** Assess metformin compared with placebo for symptom resolution during acute infection with SARS-CoV-2.

**Design, Setting, and Participants:** The ACTIV-6 platform evaluated repurposed medications for mild to moderate COVID-19. Between September 19, 2023, and May 1, 2024, 2991 participants age ≥30 years with confirmed SARS-CoV-2 infection and ≥2 COVID-19 symptoms for ≤7 days, were included at 90 US sites.

**Interventions:** Participants were randomized to receive metformin (titrated to 1500 mg daily) or placebo for 14 days.

**Main Outcomes and Measures:** The primary outcome was time to sustained recovery (3 consecutive days without COVID-19 symptoms) within 28 days of receiving study drug. Secondary outcomes included time to hospitalization or death; time to healthcare utilization (clinic visit, emergency department visit, hospitalization, or death). Safety events of special interest were hypoglycemia and lactic acidosis.

**Results:** Among 2991 participants who were randomized and received study drug, the median age was 47 years (IQR 38–58); 63.4% were female, 46.5% identified as Hispanic/Latino, and 68.3% reported ≥2 doses of a SARS-CoV-2 vaccine. Among 1443 participants who received metformin and 1548 who received placebo, differences in time to sustained recovery were not observed (adjusted hazard ratio [aHR] 0.96; 95% credible interval [CrI] 0.89–1.03; P(efficacy)=0.11). For participants enrolled during current variants, the aHR was 1.19 (95% CrI 1.05–1.34). The median time to sustained recovery was 9 days (95% confidence interval [CI] 9–10) for metformin and 10 days (95% CI 9–10) for placebo. No deaths were reported; 111 participants reported healthcare utilization: 58 in the metformin group and 53 in the placebo group (HR 1.24; 95% CrI 0.81–1.75; P(efficacy)=0.135). Seven participants who received metformin and 3 who received placebo experienced a serious adverse event over 180 days. Five participants in each group reported having hypoglycemia.

**Conclusions and Relevance:** In this randomized controlled trial, metformin was not shown to shorten the time to symptom resolution in adults with mild to moderate COVID-19. The median days to symptom resolution was numerically but not significantly lower for metformin. Safety was not a limitation in the study population.

**Trial Registration:** ClinicalTrials.gov (NCT04885530).

## INTRODUCTION

Metformin has been the first-line medication for type 2 diabetes for decades. Metformin has also been shown to exhibit *in vitro* antiviral actions in studies with dengue, hepatitis C, and rotavirus, potentially via host-directed modulation of the mammalian target of rapamycin (mTOR) pathway.^1–9^ Observational and prospective analyses have reported that prevalent metformin use was associated with less severe outcomes during acute infection with severe acute respiratory syndrome coronavirus 2 (SARS-CoV-2).^10–18^ In the COVID-OUT randomized trial of metformin versus placebo, the outcome of hospitalization / death occurred in 1.3% of the metformin group and 3.2% of the placebo group, suggesting that further study was warranted to validate these findings.^13^

As effective vaccines, natural immunity, and antivirals now make hospitalizations and deaths from coronavirus disease 2019 (COVID-19) rare, outpatient treatment for symptom resolution remains a need. The Accelerating Coronavirus Disease 2019 Therapeutic Interventions and Vaccines (ACTIV-6) platform randomized clinical trial evaluated repurposed medications in the outpatient setting.^19^ For this study, the ACTIV-6 platform sought to evaluate the effect of 14 days of immediate-release metformin, titrated to 1500 mg over 6 days, on time to sustained recovery from mild to moderate COVID-19 in non-hospitalized adults.

## METHODS

### Trial Design and Oversight

ACTIV-6 was a double-blind randomized placebo-controlled platform trial of repurposed medications in outpatient adults with mild to moderate COVID-19.^19,20^ ACTIV-6 was decentralized, allowing virtual enrollment and enrollment through health care and community settings, to improve access. The trial protocol was approved by a central institutional review board with review at each site. Informed consent was obtained from each participant. An independent data and safety monitoring committee oversaw participant safety, efficacy, and trial conduct. Reporting followed CONSORT guidelines for randomized trials.^21^

### Participants

The ACTIV-6 platform was open to enrollment from June 11, 2021, to April 29, 2024, with ∼11,000 participants randomized across 7 interventions. The metformin group enrolled participants from 90 U.S. sites between September 19, 2023 and May 1, 2024 (**Supplement**).

Eligibility criteria were confirmed at the site level and included age ≥30 years, confirmed SARS-CoV-2 infection (positive polymerase chain reaction or home-based antigen test) within 10 days, and actively experiencing ≥2 COVID-19 symptoms for ≤7 days at the time of consent. Individuals meeting the following criteria were excluded: current hospitalization for COVID-19; current or recent use (within 14 days) of metformin, insulin, or sulfonylureas; and known allergy or contraindications to metformin. Prior infection, receipt of COVID-19 vaccinations, or current use of approved or emergency use authorization therapeutics for COVID-19 were allowed.

### Randomization

Enrollment for metformin did not overlap with other agents on the platform. A random number generator randomized participants 1:1 to metformin or placebo.

### Interventions

Metformin (immediate-release tablets manufactured by Aurobindo Pharma USA Inc., East Windsor, NJ, USA) or placebo was dispensed to participants via home delivery from a centralized pharmacy. Matching placebo was available after November 9, 2023; 134 participants in the placebo group were enrolled when generic placebo was used. Participants were instructed to self-administer oral metformin (or placebo) at a dose of 500 mg per day (one 500 mg tablet) for 1 day, followed by 500 mg twice a day for 4 days, followed by 500 mg in the morning and 1000 mg in the evening for 9 days, for a 14-day course of 36 total tablets.

### Outcome Measures

The primary outcome was time to sustained recovery within 28 days, defined as the time from receipt of drug to the third of 3 consecutive days without any COVID-19 symptoms.^7,9^ This measure was selected *a priori* from the 2 possible primary outcomes of the platform, per the statistical analysis plan. The other— time to hospitalization or death—transitioned to a secondary outcome because there were not sufficient clinical outcomes in the trial overall to study those events as the primary outcome.

Pre-specified secondary outcomes included 3 time-to-event endpoints administratively censored at day 28 (number of events permitting): time to death; time to hospitalization or death; and time to first healthcare utilization (a composite of clinic visit, emergency department visit, hospitalization, or death). Additional outcomes included mean time spent unwell through day 14 and the WHO COVID Clinical Progression Scale on days 7, 14, and 28 (**Supplement**). Long COVID symptoms and patient-reported outcomes are being collected through day 180 and are not included in this report.

### Trial Procedures

Screening and eligibility information were self-reported by participants; infection status was confirmed by sites. Demographic information, medical history, use of concomitant medications, and COVID-19 symptoms were self-reported. The centralized investigational pharmacy packaged and shipped active or placebo study products to participants. Day 1 was defined based on receipt of study product as tracked by the courier. Daily assessments were sent to participants via the online REDcap electronic data capture portal during the first 14 days, regardless of symptom status.^22,23^ If participants still had symptoms on day 14, the daily assessments continued until sustained recovery or day 28. Planned remote follow-up visits occurred on days 28, 90, 120, 150, and 180.

Two safety events of interest were added for the metformin protocol: lactic acidosis or patient-reported hypoglycemia. Participants were not instructed to monitor blood sugar and the study did not send glucometers.

### Statistical Analysis Plan

Inferences about the primary outcome and exploratory analyses involving secondary outcomes were based primarily on covariate-adjusted regression modeling, supplemented by unadjusted models. Proportional hazard regression was used for time-to-event analysis, and cumulative probability ordinal regression models were used for ordinal outcomes. Differences in the mean time spent unwell were estimated with longitudinal ordinal regressions models.^24^

The primary endpoint analysis was a Bayesian proportional hazards model. The primary inferential quantity was the posterior distribution for the treatment assignment hazard ratio (HR), with a HR >1 indicating benefit. If the posterior probability of benefit exceeded 0.95 during interim or final analyses, the efficacy threshold would be met. To preserve type I error <0.05, the prior for the treatment effect parameter on the log relative hazard scale was a normal distribution centered at 0 and scaled to a standard deviation (SD) of 0.1. All other parameter priors were weakly informative, using the software default of 2.5 times the ratio of the SD of the outcome divided by the SD of the predictor variable. The study was designed to have 80% power to detect a HR of 1.2 in the primary endpoint.

The model for the primary endpoint included predictor variables: randomization assignment, age, sex, duration of symptoms prior to study drug receipt, calendar time, vaccination status, geographical location, call center indicator, and day 1 symptom severity. The proportional hazard assumption of the primary endpoint was evaluated by generating visual diagnostics such as the log-log plot and plots of time-dependent regression coefficients for each predictor in the model.

Secondary endpoints were analyzed with Bayesian regression models (either proportional hazards or proportional odds). Weakly informative priors were used for all parameters. Analyses resulting from secondary endpoints should be interpreted as exploratory given the potential for type I errors due to multiple comparisons.

All available data were used to compare metformin versus placebo, regardless of post-randomization adherence. The modified intention-to-treat (mITT) cohort comprised all participants who were randomized, who did not withdraw before study drug delivery, were not administratively withdrawn, and for whom the courier confirmed study drug delivery. Participants who opted to discontinue data collection were censored at the time of last contact, including those participants who did not complete any surveys or phone calls after study drug delivery. Missing data among covariates for both primary and secondary analyses were addressed with conditional mean imputations.

A predefined assessment of treatment effect heterogeneity (HTE) encompassed age, symptom duration, body mass index (BMI), symptom severity on day 1, calendar time (indicative of circulating SARS-CoV-2 variant), sex, and vaccination status. A proportion of participants filled out day 1 symptom surveys after taking the first dose of metformin or placebo, so a sensitivity analysis was done to remove adjusting for post-intervention symptoms or side effects.^25,26^ Metformin can cause a statistically significant, but not necessarily clinically significant, increase in diarrhea, a known side effect and also a COVID-19 symptom, soon after intake.^13^

Analyses were performed with R^10^ version 4.4 with the following primary packages: rstanarm,^11,12^ rmsb,^13^ and survival.^14^

## RESULTS

### Study Population

Of the 3214 participants randomized, 223 (6.9%) were excluded because study drug was not delivered within 7 days, or they were administratively withdrawn for not providing accurate confirmation of infection. Thus, the mITT analysis included 2991 participants, of whom 1443 (48.2%) received metformin and 1548 (51.8%) received placebo (**Figure 1**). Among the mITT participants, 5.9% confirmed receiving but taking no study medication and were not included in the safety analysis.

**Figure 1.**
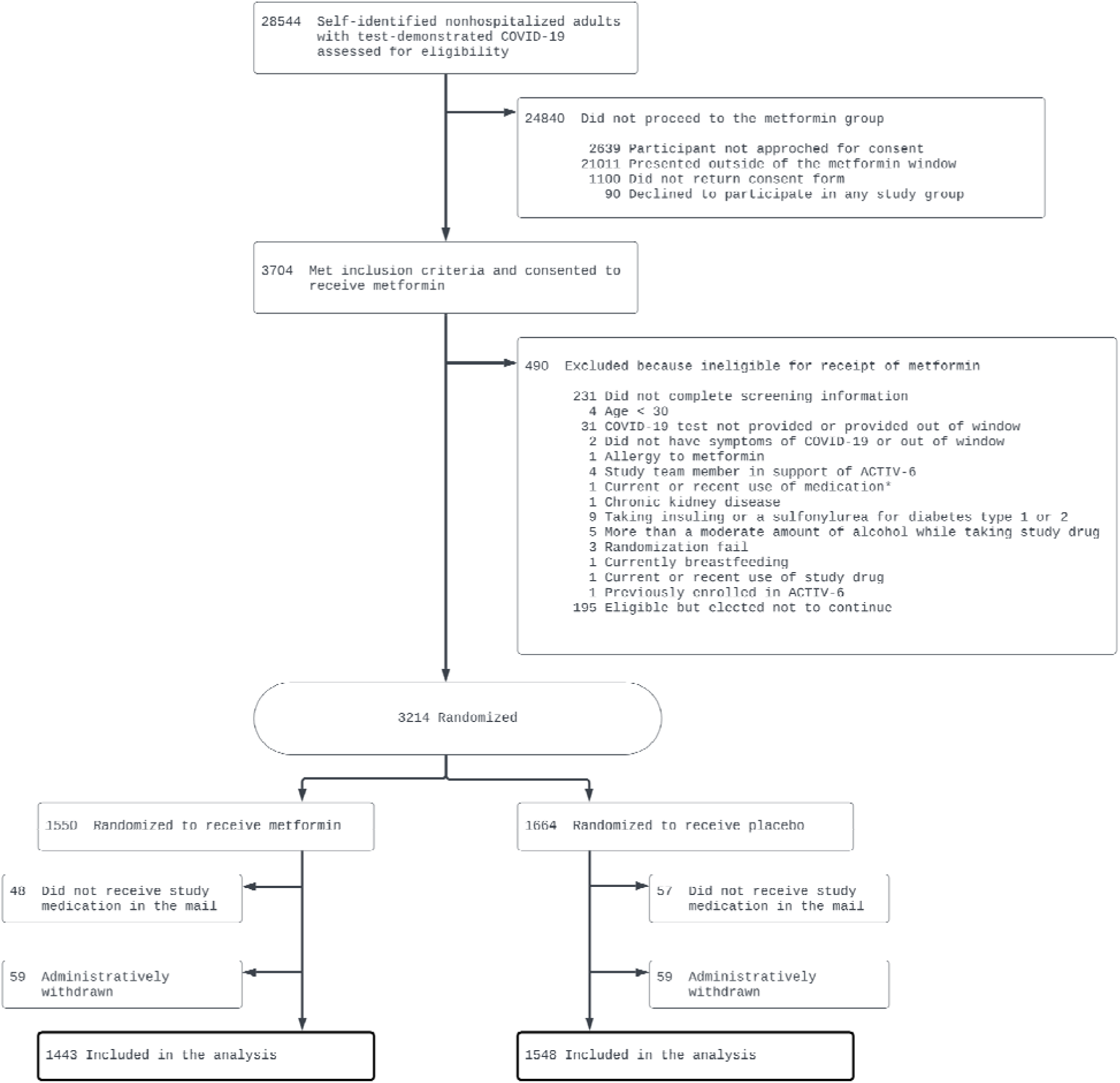
Participant flow in a trial of metformin for mild to moderate COVID-19.

The median age of the mITT population was 47 years (interquartile range [IQR] 38–58); 63.4% were female, 80.1% were White, 11.7% were Black, 0.3% were American Indian or Alaska Native, and 46.5% identified as Hispanic/Latino. Overall, 2% had diabetes, 3% had heart disease, 68.3% reported ≥2 doses of a SARS-CoV-2 vaccine, and 38.3% had obesity. Comorbidities were well balanced, though 40.1% in the metformin group and 36.6% in the placebo group had obesity (**Table 1**).

**Table 1.** Baseline participant characteristics.

| Variable | Metformin<br>(n=1443) | Placebo<br>(n=1548) |
| --- | --- | --- |
| Age, median (IQR), yrs | 47.0 (38.0-57.5) | 48.0 (37.0-58.0) |
| Age < 50 yrs, no. (%) | 798 (55.3) | 846 (54.7) |
| Sex, no. (%) |  |  |
| Female | 918 (63.62) | 977 (63.11) |
| Male | 522 (36.17) | 566 (36.56) |
| Undifferentiated | 1 (0.07) | 1 (0.06) |
| Unknown | 1 (0.07) | 2 (0.13) |
| Prefer not to answer | 1 (0.07) | 2 (0.13) |
| Race<sup>a</sup>, not mutually exclusive, no. (%) |  |  |
| American Indian or Alaska Native | 18 (1.25) | 7 (0.45) |
| Asian | 37 (2.56) | 40 (2.58) |
| Black, African American, or African | 164 (11.37) | 186 (12.02) |
| Middle Eastern or North African | 35 (2.43) | 24 (1.55) |
| Native Hawaiian or Other Pacific Islander | 4 (0.28) | 4 (0.26) |
| White | 1162 (80.53) | 1233 (79.65) |
| None of the above | 31 (2.15) | 37 (2.39) |
| Prefer not to answer | 25 (1.73) | 28 (1.81) |
| Ethnicity, no. (%) |  |  |
| Hispanic or Latino | 680 (47.12) | 712 (45.99) |
| Not Hispanic or Latino | 763 (52.88) | 836 (54.01) |
| Region<sup>b</sup>, no. (%) |  |  |
| Midwest | 295 (20.44) | 308 (19.90) |
| Northeast | 119 (8.25) | 135 (8.72) |
| South | 890 (61.68) | 942 (60.85) |
| West | 139 (9.63) | 163 (10.53) |
| Recruited via call center<sup>c</sup>, no. (%) | 41 (2.84) | 42 (2.71) |
| BMI, median (IQR), kg/m<sup>2</sup> | 28.6 (25.4-32.6) | 28.3 (25.1-31.9) |
| BMI >30 kg/m<sup>2</sup>, no./total (%) | 579/1443 (40.1) | 566/1547 (36.6) |
| Weight, median (IQR), kg | 81.6 (70.8-94.0) | 79.8 (70.3-91.2) |
| Medical history<sup>d</sup>, no./total (%) |  |  |
| High blood pressure | 282/1386 (20.35) | 316/1481 (21.34) |
| Diabetes | 31/1386 (2.24) | 27/1481 (1.82) |
| Smoker, past year | 197/1386 (14.21) | 229/1481 (15.46) |
| Asthma | 142/1386 (10.25) | 135/1481 (9.12) |
| Heart disease | 41/1386 (2.96) | 48/1481 (3.24) |
| Malignant cancer | 29/1384 (2.10) | 31/1476 (2.10) |
| COPD | 30/1386 (2.16) | 28/1481 (1.89) |
| Chronic kidney disease | 1/1386 (0.07) | 6/1481 (0.41) |
| SARS-CoV-2 vaccine status, no. (%) |  |  |
| Not vaccinated | 463 (32.09) | 480 (31.01) |
| Vaccinated, 1 dose | 2 (0.14) | 2 (0.13) |
| Vaccinated, 2+ doses | 978 (67.78) | 1066 (68.86) |
| Days between symptom onset and receipt of drug, median (IQR) | 5 (4-7)<sup>e</sup> | 5 (4-7)<sup>f</sup> |
| Days between symptom onset and enrollment, median (IQR) | 3 (2-5)<sup>e</sup> | 3 (2-5)<sup>f</sup> |

| Symptom burden on study day 1<sup>g</sup>, no./No. (%) |  |  |
| --- | --- | --- |
| None | 68/1322 (5.1) | 66/1388 (4.8) |
| Mild | 610/1322 (46.1) | 660/1388 (47.6) |
| Moderate | 610/1322 (46.1) | 617/1388 (44.5) |
| Severe | 34/1322 (2.6) | 45/1388 (3.2) |
| COVID-19 medications, no. (%) |  |  |
| Remdesivir | 1 (0.07) | 0 (0.00) |
| Nirmatrelvir + ritonavir | 204 (14.14) | 229 (14.79) |
| Monoclonal antibodies | 31 (2.15) | 43 (2.78) |
| Molnupiravir | 10 (0.69) | 13 (0.84) |

Abbreviations: BMI, body mass index (calculated as weight in kilograms divided by height in meters squared); COPD, chronic obstructive pulmonary disease.
<sup>a</sup>Participants were presented with each of these options and may have selected any combination of the race descriptors, including “none of the above” and “prefer not to answer.” Consequently, the sums of counts over all categories do not match the column totals.
<sup>b</sup>The following state groups define each region. Northeast includes Connecticut, Maine, Massachusetts, New Hampshire, Rhode Island, Vermont, New Jersey, New York, and Pennsylvania; Midwest includes Indiana, Illinois, Michigan, Ohio, Wisconsin, Iowa, Kansas, Minnesota, Missouri, Nebraska, North Dakota, and South Dakota; South includes Delaware, District of Columbia, Florida, Georgia, Maryland, North Carolina, South Carolina, Virginia, West Virginia, Alabama, Kentucky, Mississippi, Tennessee, Arkansas, Louisiana, Oklahoma, and Texas; West includes Arizona, Colorado, Idaho, New Mexico, Montana, Utah, Nevada, Wyoming, Alaska, California, Hawaii, Oregon, and Washington.
<sup>c</sup>Patients may have alternatively been recruited at local clinical sites.
<sup>d</sup>Medical history was provided by participants responding to the following prompts: “Has a doctor told you that you have any of the following?” “Have you ever experienced any of the following? (select all that apply),” and “Have you ever smoked tobacco products?”
<sup>e</sup>Among 1411 participants;
<sup>f</sup>Among 1511 participants
<sup>i</sup>Each day, participants were asked, “Please choose the response that best describes the severity of your COVID-19 symptoms today.”

Study drug was delivered within a median of 5 days of symptom onset (IQR 4–7 days). The distribution of time between COVID-19 symptoms and study drug delivery is reported in **eFigure 1**. Participants who achieved 3 days without symptoms on or before study drug delivery were not excluded. Of the 2710 participants who submitted symptom surveys on the day of study drug delivery, 5% reported no symptoms, 47% reported mild symptoms, 45% reported moderate symptoms, and 3% reported severe symptoms (**Table 1**, **eTable 1**).

### Primary Outcome

Overall, 87/2991(3%) did not provide any follow-up responses beyond study day 1 and an additional 152/2991 (5%) were censored due to partial follow-up responses. Differences in time to sustained recovery were not observed in either the covariate-adjusted regression models (**Table 2**) or unadjusted Kaplan-Meier curves (**Figure 2**). The median time to sustained recovery was 9 days (95% CI 9–10) in the metformin group and 10 days (95% CI 9–10) in the placebo group. The posterior probability for benefit was 0.11, with an adjusted hazard ratio (aHR) of 0.95 (95% credible interval [CrI] 0.89–1.03). In the sensitivity analysis that adjusted for symptoms at randomization to remove adjusting for post-intervention symptoms or side effects, the aHR was 0.98 (95% CrI 0.91–1.05). Sensitivity analyses based on different patterns of missing symptom data yielded similar estimates of the treatment effect (**eFigure 2**).

**Figure 2.**
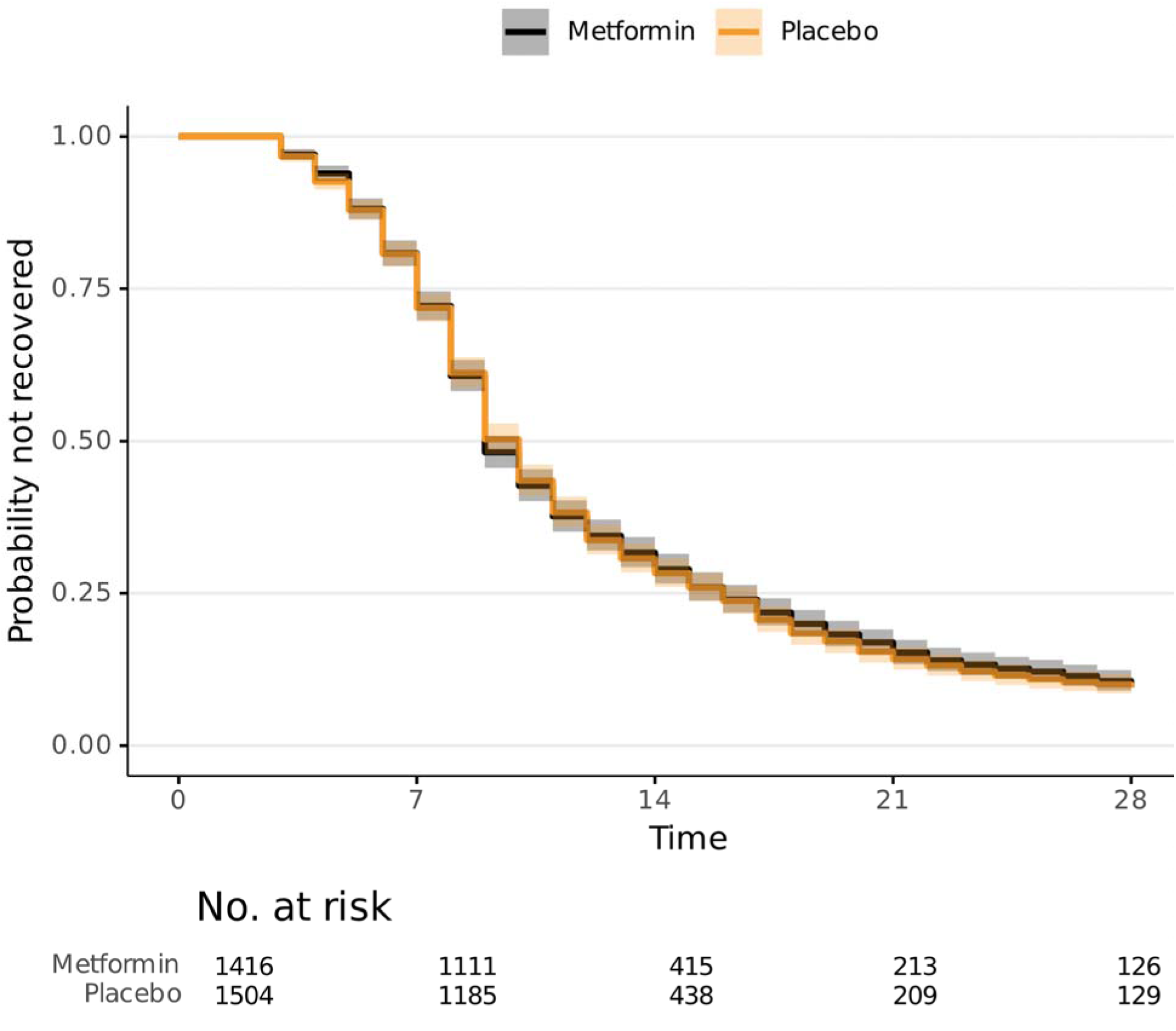
Primary outcome of time to sustained recovery. Sustained recovery was defined as the third of 3 consecutive days without symptoms. Seventy-one participants were censored for complete nonresponse and an additional 168 after partial response, 239/2,991, 8%; all others were followed up until recovery, death, or the end of short-term 28-day follow-up. Median time to sustained recovery was 9 days (95% CI, 9-10 days) in the metformin group and 10 days (95% CI, 9-10 days) in the placebo group. Shading denotes the pointwise 95% confidence intervals.

**Table 2.**
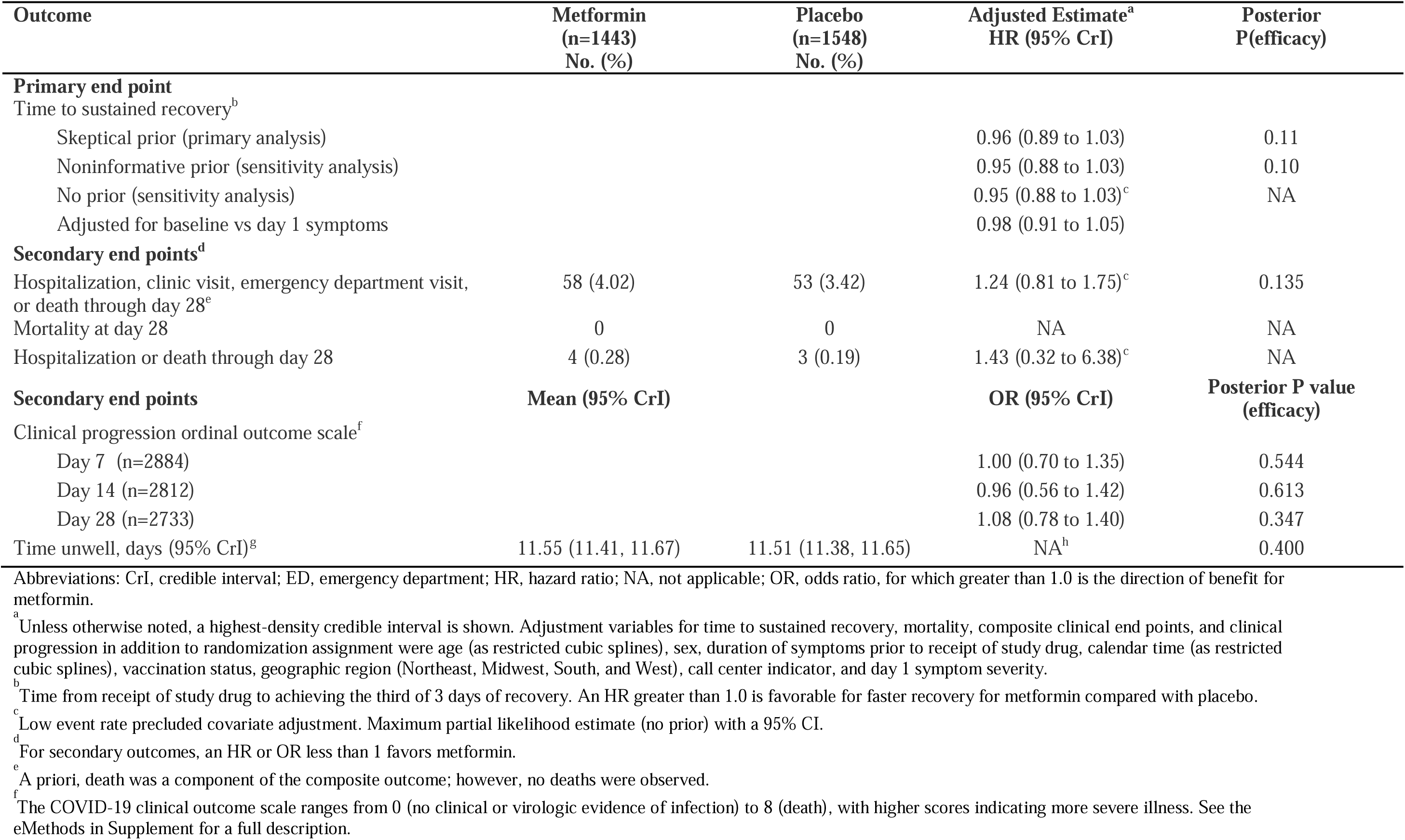
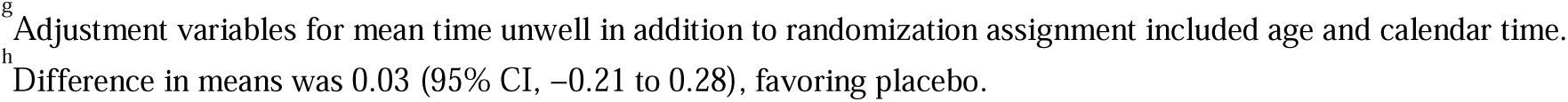
Primary and secondary outcomes.

| Outcome | Metformin<br>(n=1443)<br>No. (%) | Placebo<br>(n=1548)<br>No. (%) | Adjusted Estimate<sup>a</sup><br>HR (95% CrI) | Posterior<br>P(efficacy) |
| --- | --- | --- | --- | --- |
| **Primary end point** |  |  |  |  |
| Time to sustained recovery<sup>b</sup> |  |  |  |  |
| Skeptical prior (primary analysis) |  |  | 0.96 (0.89 to 1.03) | 0.11 |
| Noninformative prior (sensitivity analysis) |  |  | 0.95 (0.88 to 1.03) | 0.10 |
| No prior (sensitivity analysis) |  |  | 0.95 (0.88 to 1.03)<sup>c</sup> | NA |
| Adjusted for baseline vs day 1 symptoms |  |  | 0.98 (0.91 to 1.05) |  |
| **Secondary end points<sup>d</sup>** |  |  |  |  |
| Hospitalization, clinic visit, emergency department visit, or death through day 28<sup>e</sup> | 58 (4.02) | 53 (3.42) | 1.24 (0.81 to 1.75)<sup>c</sup> | 0.135 |
| Mortality at day 28 | 0 | 0 | NA | NA |
| Hospitalization or death through day 28 | 4 (0.28) | 3 (0.19) | 1.43 (0.32 to 6.38)<sup>c</sup> | NA |
| **Secondary end points** | **Mean (95% CrI)** |  | **OR (95% CrI)** | **Posterior P value**<br>**(efficacy)** |
| Clinical progression ordinal outcome scale<sup>f</sup> |  |  |  |  |
| Day 7 (n=2884) |  |  | 1.00 (0.70 to 1.35) | 0.544 |
| Day 14 (n=2812) |  |  | 0.96 (0.56 to 1.42) | 0.613 |
| Day 28 (n=2733) |  |  | 1.08 (0.78 to 1.40) | 0.347 |
| Time unwell, days (95% CrI)<sup>g</sup> | 11.55 (11.41, 11.67) | 11.51 (11.38, 11.65) | NA<sup>h</sup> | 0.400 |

Abbreviations: CrI, credible interval; ED, emergency department; HR, hazard ratio; NA, not applicable; OR, odds ratio, for which greater than 1.0 is the direction of benefit for metformin.
<sup>a</sup> Unless otherwise noted, a highest-density credible interval is shown. Adjustment variables for time to sustained recovery, mortality, composite clinical end points, and clinical progression in addition to randomization assignment were age (as restricted cubic splines), sex, duration of symptoms prior to receipt of study drug, calendar time (as restricted cubic splines), vaccination status, geographic region (Northeast, Midwest, South, and West), call center indicator, and day 1 symptom severity.
<sup>b</sup> Time from receipt of study drug to achieving the third of 3 days of recovery. An HR greater than 1.0 is favorable for faster recovery for metformin compared with placebo.
<sup>c</sup> Low event rate precluded covariate adjustment. Maximum partial likelihood estimate (no prior) with a 95% CI.
<sup>d</sup> For secondary outcomes, an HR or OR less than 1 favors metformin.
<sup>e</sup> A priori, death was a component of the composite outcome; however, no deaths were observed.
<sup>f</sup> The COVID-19 clinical outcome scale ranges from 0 (no clinical or virologic evidence of infection) to 8 (death), with higher scores indicating more severe illness. See the eMethods in Supplement for a full description.
<sup>g</sup> Adjustment variables for mean time unwell in addition to randomization assignment included age and calendar time. <sup>h</sup> Difference in means was 0.03 (95% CI, -0.21 to 0.28), favoring placebo.

### Secondary Outcomes

No deaths were observed in this study. A total of 7/2991 participants (0.23%) were hospitalized (**Table 2, eFigure 3**). There were 58 (4.0%) participants in the metformin group and 53 (3.4%) in the placebo group who reported a hospital admission, emergency department visit, or clinic visit (**Table 2**, **eFigure 3**), aHR for the composite healthcare encounter outcome was 1.24 (95% CrI 0.81–1.75) with a posterior probability of efficacy of 0.135.

By day 7, 163/2991 (5.45%) reported limitation on activities. Because 94% of responding participants reported no limitations in activity on the COVID clinical progression scale, this endpoint did not meet prespecified thresholds for beneficial treatment effect (**eFigure 4**). Likewise, the mean time unwell was similar between the metformin and placebo groups, 11.6 days (CrI 11.4–11.7) versus 11.5 days (CrI 11.4–11.7), respectively, (difference 0.03, 95% CrI −0.21 to 0.28; P[efficacy]=0.40) (**Table 2**, **Figure 3).**

**Figure 3A.**
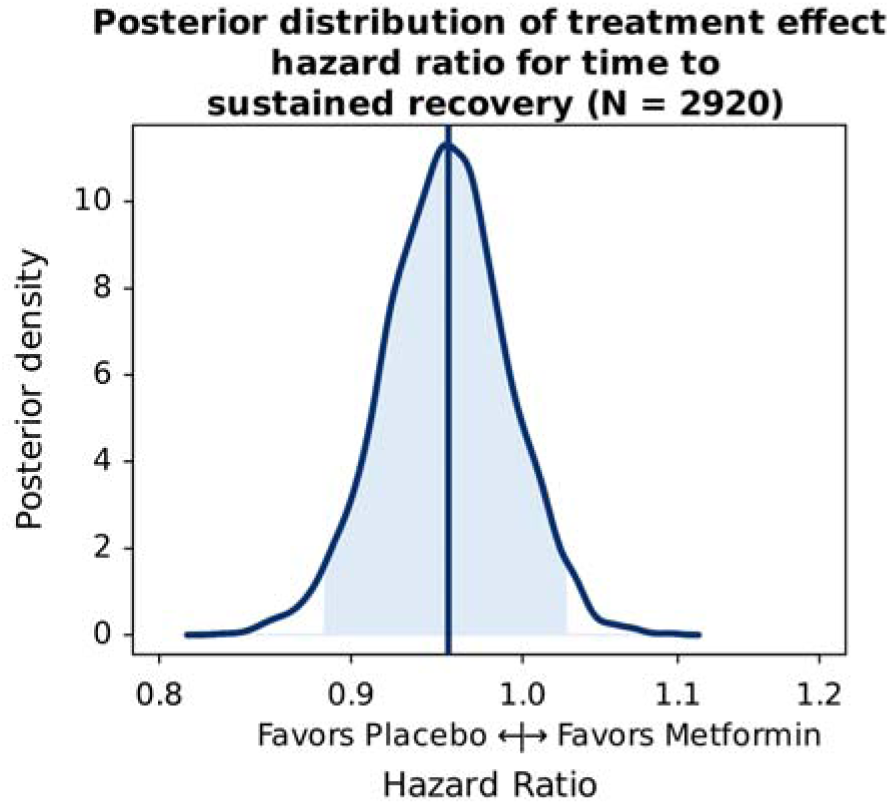
Posterior distribution of treatment effect hazard ratio for time to sustained recovery. Posterior density is the relative likelihood of posterior probability distribution. Outcomes with higher posterior density are more likely than outcomes with lower posterior density. Blue density lines represent kernel density estimates constructed from posterior draws; vertical lines, estimated means of the posterior distribution; shading, the 95% credible interval. Posterior density plots of all the covariates in the primary outcome model are shown in eFigure 8 in Supplement.

**Figure 3B.**
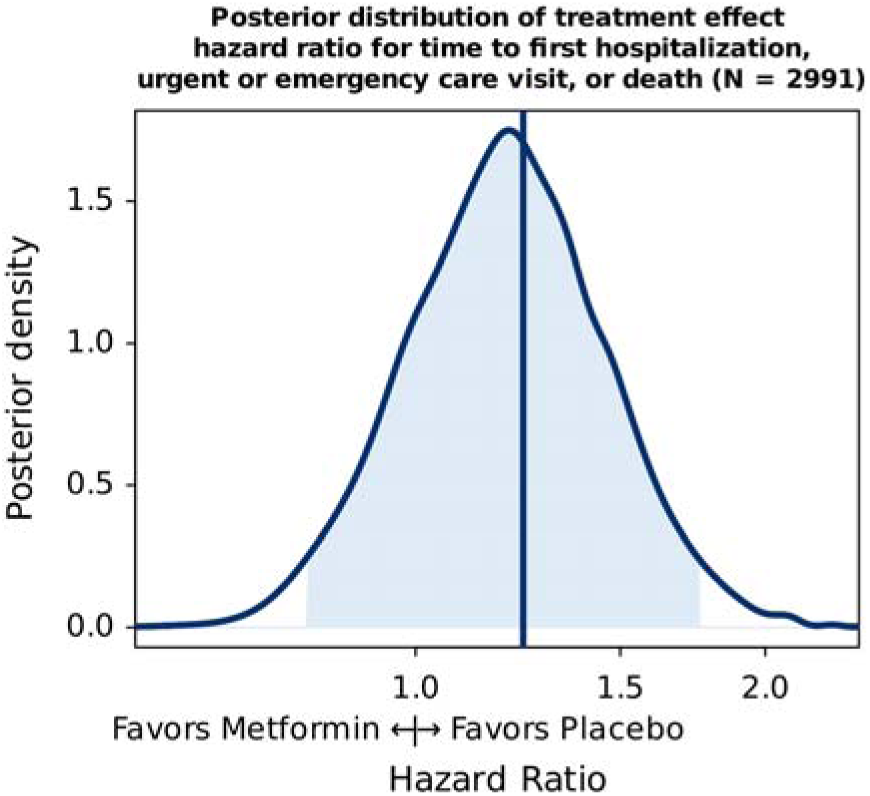
Posterior distribution of treatment effect hazard ratio for time to first hospitalization, urgent or emergency care visit, or death. Posterior density is the relative likelihood of posterior probability distribution. Outcomes with higher posterior density are more likely than outcomes with lower posterior density. Blue density lines represent kernel density estimates constructed from posterior draws; vertical lines, estimated means of the posterior distribution; shading, the 95% credible interval. Posterior density plots of all the covariates in the primary outcome model are shown in eFigure 8 in Supplement.

**Figure 3C.**
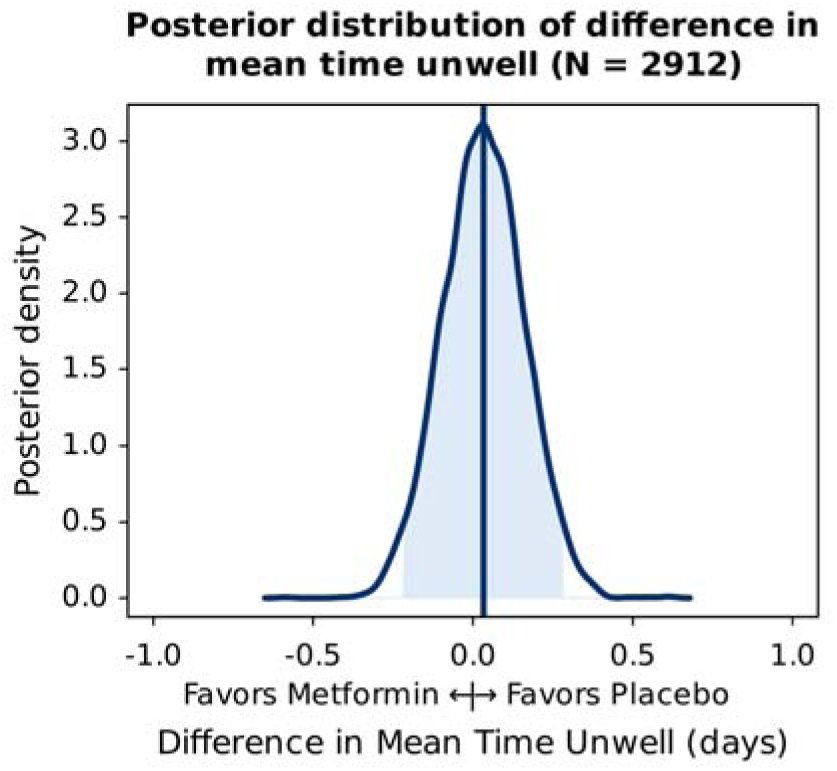
Posterior distribution of difference in mean time unwell. Posterior density is the relative likelihood of posterior probability distribution. Outcomes with higher posterior density are more likely than outcomes with lower posterior density. Blue density lines represent kernel density estimates constructed from posterior draws; vertical lines, estimated means of the posterior distribution; shading, the 95% credible interval. Posterior density plots of all the covariates in the primary outcome model are shown in eFigure 8 in Supplement.

### Adverse Events and Events of Special Interest

The safety sample for collecting serious adverse events was the 2815/2991 (94.1%) of participants who reported taking study medication at least once. The incidence of serious events over 180 days was low (11 total events within 10/2825 [0.35%] of participants), with 8 events (7 participants) in the metformin group and 3 events (3 participants) in the placebo group. There were no episodes of lactic acidosis and 5 episodes of patient-reported hypoglycemia in each treatment group (**eTable 2**).

### Heterogeneity of Treatment Effect Analyses

In participants with no symptoms on day 1, the aHR for time to symptom recovery was 0.40 (95% CI 0.28–0.58; P<0.01). The interaction model created a calendar time category for those enrolled after March 15, 2024, which is when the FLiRT variants became dominant. In the 349 (12%) participants enrolled after March 15, 2024, the aHR for time to recovery was 1.19 (95% CI 1.05–1.34; P=0.82) (**eFigure 5**).

The primary analysis was stratified by day 1 symptom severity and timing of drug delivery relative to symptom onset. In those with no symptoms on day 1, the curves separated in the direction of benefit for placebo; in those with moderate and severe symptoms, the curves separated in the direction of benefit for metformin (**eFigures 6, 7**).

Other baseline characteristics of interest, including history of previous infection and body mass index (BMI), were assessed in post-hoc exploratory analyses. Overall, 71 participants did not provide follow-up information, so 44/2920 (1.5%) had an acute healthcare encounter by day 28: 1.4% in the metformin group and 1.6% in the placebo group. In those with no reported prior infection, 0.99% experienced an acute healthcare encounter in the metformin group and 1.32% in the placebo group (**eTable 3**). There was no indication that participants with a BMI <25 kg/m^2^ had a longer time to recovery or more healthcare events (**eTable 3**).

## DISCUSSION

Among outpatient adults with mild to moderate COVID-19, treatment with immediate-release metformin did not improve time to sustained recovery compared with placebo. Other studies that investigated metformin as an outpatient treatment for SARS-CoV-2 infection evaluated severe clinical events and had limitations: the TOGETHER trial (n=421) had exposure misclassification, but a partially corrected analysis showed similar results for preventing emergency department visits and hospitalizations as COVID-OUT^27–29^ and the COVID-OUT trial (n=1323) had outcome misclassification, with the primary outcome driven by the sensitive but non-specific component of the composite (1 oxygen reading <94% on a home oximeter).^13^ Of those randomized trials, COVID-OUT is the most generalizable to the current state of the pandemic because it enrolled vaccinated individuals and was enrolling after the Omicron wave started. The ACTIV-6 study of metformin has further relevance as it is the first trial to allow individuals with a reported history of prior infection to enroll.

The incidence of severe outcomes was less than one-quarter of a percent in the ACTIV-6 metformin arm. Such a low incidence means a trial would need to enroll tens of thousands of participants to test whether treatment prevents hospitalizations and deaths, which is why a symptom recovery outcome may be more relevant at this point in the pandemic. However, antivirals developed for SARS-CoV-2 have not reduced symptoms during acute infection.^30,31^ Additionally, most participants were enrolled between December 2023 and March 2024, when the JN-1 variant dominated.^32^ JN-1 had higher infectivity but lower symptom and disease burden than previous variants.^33,34^ Most participants achieved symptom recovery by day 28, and outcomes with high incidence can make it challenging to detect intervention differences.^13,35^ A portion of the trial was enrolled after the FLiRT variants began, and these variants include mutations similar to those in earlier Omicron subvariants.^36,37^

Strengths of this study include high ethnic diversity with 47% of participants identifying as Hispanic/Latino ethnicity, a result of concerted efforts to translate documents and onboard sites already engaged with diverse communities. Metformin has been studied in hundreds of clinical trials in patients with some degree of chronic disease or risk for disease, such as diabetes, diabetes risk factors, and cancer. This is the first phase 3 trial to randomize adults without chronic disease or disease risk to metformin, and there were no episodes of low blood sugar or lactic acidosis, despite a rapid dose escalation to 1500 mg in just 6 days.

### Limitations

This study does have limitations. Due to the decentralized nature of ACTIV-6, time from symptom onset to receipt of study drug is approximately 2 days longer than time from symptom onset to enrollment. Trials of antivirals assessing outpatient treatment of SARS-CoV-2, such as nirmatrelvir, simnotrelvir, ensitrelvir, and molnupiravir, restricted eligibility or the primary analysis to participants enrolled within 3 days of COVID-19 symptom onset.^30,31,38^ ACTIV-6 inclusion criteria allowed enrollment of participants with up to 7 days of symptoms, in part due to various mechanisms of action of the drugs tested on the platform. However, this resulted in a relatively small number of participants in this study (<21%) receiving drug within 3 days of symptom onset. Because emerging data from *in vivo and in vitro* studies have shown that metformin has antiviral activity against SARS-Co-V-2, a time-dependent treatment effect is expected.^14,39–43^ An additional limitation for a primary outcome of symptom resolution is that 5% of the mITT participants had no symptoms on the day of study drug delivery. Prespecified and exploratory results are directionally consistent with hypotheses about benefit with earlier initiation, consistent with other studies assessing earlier initiation of metformin, but underpowered subgroups cannot be over-interpreted.^13,44,45^ This study also had a higher proportion of post-randomization exclusions compared with earlier studies, and the placebo group is 7% larger than the metformin group. Information on effect modifiers such as time of study drug initiation and previous infection could not always be captured, and nonresponse for primary and secondary outcome measures is less than 10% but may introduce some bias.

## Conclusions

Among low-risk outpatient adults with mild to moderate COVID-19, metformin did not shorten the time to symptom recovery compared with placebo. The median days to symptom resolution was numerically but not significantly lower for metformin than placebo. Emergency room visits or hospitalizations were too infrequent to draw conclusions between the two study groups. There were no episodes of lactic acidosis or other significant safety concerns in this trial of metformin in adults without diabetes or diabetes risk factors such as overweight and obesity.

## Data Availability

ACTIV-6 is a platform trial using shared placebos. On completion of the platform trial, when there is no risk of unblinding across study arms, the data will be made publicly available by depositing it in an approved data repository such as the NHLBI BioData Catalyst.

## Acknowledgments

We thank George J. Hanna, MD, of the Biomedical Advanced Research and Development Authority, for his roles in trial implementation and operations. We also thank the ACTIV-6 Data Monitoring Committee, Clinical Events Committee, and Stakeholder Advisory Committee Members (listed below) for their contributions.

Data Monitoring Committee: Clyde Yancy, MD, MSc, Northwestern University Feinberg School of Medicine; Adaora Adimora, MD, University of North Carolina, Chapel Hill; Susan Ellenberg, PhD, University of Pennsylvania; Kaleab Abebe, PhD, University of Pittsburgh; Arthur Kim, MD, Massachusetts General Hospital; John D. Lantos, MD, Children’s Mercy Hospital; Jennifer Silvey-Cason, Participant representative; Frank Rockhold, PhD, Duke Clinical Research Institute; Sean O’Brien, PhD, Duke Clinical Research Institute; Frank Harrell, PhD, Vanderbilt University Medical Center; Zhen Huang, MS, Duke Clinical Research Institute.

Clinical Events Committee: Renato Lopes, MD, PhD, MHS, W. Schuyler Jones, MD, Antonio Gutierrez, MD, Robert Harrison, MD, David Kong, MD, Robert McGarrah, MD, Michelle Kelsey, MD, Konstantin Krychtiuk, MD, Vishal Rao, MD all of the Duke Clinical Research Institute, Duke University School of Medicine.

Stakeholder Advisory Committee: Megan E. Hamm, Kathleen McTigue, Kirk T. Phillips, Andrew Vasey, Talethia Edwards, Danielle Nelson, Greg Merritt, Jeannie Nguyen, Josh Denson, Jonathan Arnold, Matthew W. McCarthy, Florence Thicklin.

Elizabeth E.S. Cook of the Duke Clinical Research Institute provided editorial support.

## Author Contributions

Drs Naggie, Hernandez, and Lindsell had full access to all the blinded data in the study. Dr Stewart was provided curated study data and takes responsibility for the integrity of the data analysis. All authors contributed to the drafting and review of the manuscript and agreed to submit for publication.

## Disclosures

**Bramante:** Reports grants from NIH NIDDK during the conduct of the study; Grants from NIH outside the submitted work.

**Stewart:** Reports grants from NIH NCATS during the conduct of the study; Grants from NIH outside the submitted work.

**Boulware:** Reports grants from NIH during the conduct of the study.

**McCarthy:** Nothing to report.

**Gao:** Nothing to report.

**Rothman:** Reports grants from NIH, PCORI, AHRQ, CDC, CardioHealth Alliance during the conduct of the study. Spouse owns stock in Moderna unrelated to the current work.

**Mourad:** Nothing to report.

**Thicklin:** Nothing to report.

**Cohen:** Nothing to report.

**Garcia del Sol:** Nothing to report.

**Ruiz-Unger:** Nothing to report.

**Shah:** Nothing to report.

**Mehta:** Nothing to report.

**Cardona:** Nothing to report.

**Scott:** Nothing to report.

**Ginde:** Reports grants from NIH during the conduct of the study; Grants from NIH, CDC, DoD, AbbVie (investigator-initiated), and Faron Pharmaceuticals (investigator-initiated) outside the submitted work.

**Castro:** Reports institutional grant funding from NIH, ALA, PCORI, AstraZeneca, GSK, Novartis, Pulmatrix, Sanofi-Aventis, Shionogi; Speaker/Consultant fees from Grant Funding, Genentech, Teva, Sanofi-Aventis; Consultant fees from Merck, Novartis, Arrowhead, OM Pharma, Allakos; Speaker honorarium from Amgen, AstraZeneca, GSK, Regeneron; Royalties from Elsevier all outside the submitted work.

**Jayaweera:** Reports grants from NCATS PI-Ralph Sacco during the conduct of the study; Grants from Gilead, Pfizer, Janssen, and Viiv; Consulting fees from Theratechnologies outside the submitted work.

**Sulkowski:** Reports advisory board fees from AbbVie, Gilead, GSK, Atea, Antios, Precision Bio, Viiv, and Virion; Institutional grants from Janssen outside the submitted work.

**Gentile:** Reports personal fees from Duke University for protocol development and oversight during the conduct of the study; grants from NIH outside the submitted work.

**McTigue:** Reports grants from NIH Research Subcontract to the University of Pittsburgh during the conduct of the study; Research contract to the University of Pittsburgh from Pfizer, and Janssen outside the submitted work.

**Felker:** Reports institutional research grants from NIH during the conduct of the study and from Novartis outside the submitted work.

**Collins:** Reports grant funding from NHLBI and personal fees from Vir Biotechnology during the conduct of the study.

**Dunsmore:** Nothing to report.

**Adam:** Reports other from US Government Funding through Operation Warp Speed during the conduct of the study.

**Lindsell:** Reports institutional grants from NCATS during the conduct of the study; Institutional grants from NIH, CDC, and DoD; Contract with institution for research services from Endpoint Health, bioMerieux, Entegrion Inc, Abbvie, and Astra Zeneca, Biomeme, and Novartis outside the submitted work; Dr Lindsell has a patent for risk stratification in sepsis and septic shock issued to Cincinnati Children’s Hospital Medical Center.

**Hernandez:** Reports grants from American Regent, Amgen, Boehringer Ingelheim, Merck, Verily, Somologic, and Pfizer; Personal fees from AstraZeneca, Boston Scientific, Cytokinetics, Bristol Myers Squibb, and Merck outside the submitted work.

**Naggie:** Reports grants from NIH, the sponsor for this study, during the conduct of the study; Institutional research grants from Gilead Sciences, AbbVie; Consulting fees from Pardes Biosciences; Scientific advisor/Stock options from Vir Biotechnology; Consulting with no financial payment from Silverback Therapeutics; DSMB fees from Personal Health Insights, Inc; Event adjudication committee fees from BMS/PRA outside the submitted work.

## Funding/Support

ACTIV-6 is funded by the National Center for Advancing Translational Sciences (NCATS) (3U24TR001608-06S1). Additional support for this study was provided by the Office of the Assistant Secretary for Preparedness and Response, Biomedical Advanced Research and Development Authority (Contract No.75A50122C00037). The Vanderbilt University Medical Center Clinical and Translational Science Award from NCATS (UL1TR002243) supported the REDCap infrastructure.

## Role of the Sponsor

NCATS participated in the design and conduct of the study; collection, management, analysis, and interpretation of the data; preparation, review, or approval of the manuscript; and decision to submit the manuscript for publication.

## Data Sharing Statement

ACTIV-6 is a platform trial using shared placebos. On completion of the platform trial, when there is no risk of unblinding across study arms, the data will be made publicly available by depositing it in an approved data repository such as NHLBI’s BioData Catalyst.

## Notes

### Clinical Trial

NCT04885530

### Author Declarations

WCG IRB gave ethical approval for this work.

