## Supplement for "Metformin on Time to Sustained Recovery in Adults with COVID-19: The ACTIV-6 Randomized Clinical Trial"

**Online-only Supplement**

**ACTIV-6 Executive Committee**

Adrian F. Hernandez, Duke Clinical Research Institute (Clinical Coordinating Center PI)

Susanna Naggie, Duke Clinical Research Institute (Clinical Coordinating Center Co-PI)

G. Michael Felker, Duke Clinical Research Institute (Medical Monitor, blinded)

Allison DeLong, Duke Clinical Research Institute

Rhonda Wilder, Duke Clinical Research Institute

Christopher J. Lindsell, Duke Clinical Research Institute (Data Coordinating Center Executive Director)

Russell L. Rothman, Vanderbilt University Medical Center (Data Coordinating Center PI)

Thomas G. Stewart, University of Virginia

Sean Collins, Vanderbilt University Medical Center

Sarah Dunsmore, National Center for Advancing Translational Sciences

Stacey Adam, Foundation for the National Institutes of Health

David R. Boulware, University of Minnesota

Carolyn Bramante, University of Minnesota

Florence Thicklin, Stakeholder Advisory Committee

Matthew William McCarthy, Weill Cornell Medicine, Stakeholder Advisory Committee

George Hanna, Biomedical Advanced Research and Development Authority

**ACTIV-6 Protocol Oversight Committee**

David Boulware, University of Minnesota, POC Co-Chair

Adrian F. Hernandez, Duke Clinical Research Institute (Clinical Coordinating Center PI)

Susanna Naggie, Duke Clinical Research Institute (Clinical Coordinating Center co-PI)

G. Michael Felker, Duke Clinical Research Institute (Medical Monitor, blinded)

Allison DeLong, Duke Clinical Research Institute

Rhonda Wilder, Duke Clinical Research Institute

Christopher J. Lindsell, Duke Clinical Research Institute (Data Coordinating Center Executive Director)

Russell L. Rothman, Vanderbilt University Medical Center (Data Coordinating Center PI)

Thomas G. Stewart, University of Virginia

Sean Collins, Vanderbilt University Medical Center

Sarah Dunsmore, National Center for Advancing Translational Sciences

Stacey Adam, Foundation for the National Institutes of Health

Florence Thicklin, Stakeholder Advisory Committee

Matthew William McCarthy, Weill Cornell Medicine, Stakeholder Advisory Committee

George Hanna, Biomedical Advanced Research and Development Authority

Adit Ginde, University of Colorado Denver – Anschutz

Mario Castro, University of Kansas Medical Center

Dushyantha Jayaweera, University of Miami

Mark Sulkowski, John Hopkins University

Nina Gentile, Lewis Katz School of Medicine at Temple University

Kathleen McTigue, University of Pittsburgh Medical Center

Kim Marschhauser, PCORI

**ACTIV-6 Clinical Trial Team**

Adrian F. Hernandez, Duke Clinical Research Institute (Clinical Coordinating Center PI)

Susanna Naggie, Duke Clinical Research Institute (Clinical Coordinating Center Co-PI)

G. Michael Felker, Duke Clinical Research Institute (Medical Monitor, blinded)

Allison DeLong, Duke Clinical Research Institute

Rhonda Wilder, Duke Clinical Research Institute

Christopher J. Lindsell, Duke Clinical Research Institute (Data Coordinating Center Executive Director)

Sean Collins, Vanderbilt University Medical Center (Data Coordinating Center PI)

Thomas G. Stewart, University of Virginia

**ACTIV-6 Clinical Coordinating Center**

Adrian F. Hernandez, Duke Clinical Research Institute (Clinical Coordinating Center PI)

Susanna Naggie, Duke Clinical Research Institute (Clinical Coordinating Center Co-PI)

G. Michael Felker, Duke Clinical Research Institute (Medical Monitor, blinded)

Allison DeLong, Duke Clinical Research Institute

Rhonda Wilder, Duke Clinical Research Institute

Ryan Fraser, Duke Clinical Research Institute

Mark Ward, Duke Clinical Research Institute

Sarah Weaver, Duke Clinical Research Institute

M. Patricia McAdams, Duke Clinical Research Institute

Kayla Korzekwinski, Duke Clinical Research Institute

Martina Oyelakin, Duke Clinical Research Institute

Samantha Dockery, Duke Clinical Research Institute

Rodney Adkins, Duke Clinical Research Institute

Mathew Crow, Duke Clinical Research Institute

Shanee Light, Duke Clinical Research Institute

Renee Morris, Duke Clinical Research Institute

Erin Nowell, Duke Clinical Research Institute

Kadie Wells, Duke Clinical Research Institute

Alicia Herbert, Duke Clinical Research Institute

Allegra Stone, Duke Clinical Research Institute

Heather Heavlin, Duke Clinical Research Institute

Linley Brown, Duke Clinical Research Institute

Shelley Brunson, Duke Clinical Research Institute

Tina Harding, Duke Clinical Research Institute

Amanda Harrington, Duke Clinical Research Institute

Meaghan Beauchaine, Duke Clinical Research Institute

Kelly Lindblom, Duke Clinical Research Institute

Andrea Burns, Duke Clinical Research Institute

Ahmad Mourad, Duke Clinical Research Institute

**ACTIV-6 Stakeholder Advisory Committee**

Megan E. Hamm

Kathleen McTigue

Kirk T. Phillips

Andrew Vasey

Talethia Edwards

Danielle Nelson

Greg Merritt

Jeannie Nguyen

Josh Denson

Jonathan Arnold

Matthew McCarthy

Florence Thicklin

**ACTIV-6 Independent Data Monitoring Committee**

**Voting Members:**

Clyde Yancy, Northwestern University Feinberg School of Medicine (Chair)

Adaora Adimora, University of North Carolina, Chapel Hill (Vice-chair)

Susan Ellenberg, University of Pennsylvania

Kaleab Abebe, University of Pittsburgh

Arthur Kim, Massachusetts General Hospital

John D. Lantos, Children’s Mercy Hospital

Jennifer Silvey-Cason, Participant representative

**Statistical Data Analysis Center:**

Frank Rockhold, Duke Clinical Research Institute (Lead faculty statistician)

Sean O’Brien, Duke Clinical Research Institute (Faculty statistician)

Frank Harrell, Vanderbilt University Medical Center (DCC-SDAC Faculty Liaison)

Zhen Huang, Duke Clinical Research Institute (Lead statistician)

Hayley Nemeth, Duke Clinical Research Institute

**ACTIV-6 Clinical Events Classification Committee**

Renato Lopes, (CEC PI)

**Faculty Reviewers:** W. Schuyler Jones, Antonio Gutierrez, Robert Harrison, David Kong, Robert McGarrah, Michelle Kelsey, Brad Kolls, Cina Sasannejad, Rajendra (Raj) Mehta,

**Fellow Reviewers:** Mark Kittipibul

**ACTIV-6 Data Coordinating Center**

David Aamodt, Vanderbilt University Medical Center

Deborah Clark, Vanderbilt University Medical Center

Jess Collins, Vanderbilt University Medical Center

Sean Collins, Vanderbilt University Medical Center (Data Coordinating Center PI)

Sheri Dixon, Vanderbilt University Medical Center

Yue Gao, Vanderbilt University Medical Center

John Graves, Vanderbilt University Medical Center

James Grindstaff, Vanderbilt University Medical Center

Frank Harrell, Vanderbilt University Medical Center (DCC-SDAC Faculty Liaison)

Jessica Lai, Vanderbilt University Medical Center

Vicky Liao, Vanderbilt University Medical Center

Christopher J. Lindsell, Duke Clinical Research Institute (Data Coordinating Center Executive Director)

Itzel Lopez, Vanderbilt University Medical Center

Elizabeth Manis, Vanderbilt University Medical Center

Kalley Mankowski, Vanderbilt University Medical Center

Jessica Marlin, Vanderbilt University Medical Center

Alyssa Merkel, Vanderbilt University Medical Center

Sam Nwosu, Vanderbilt University Medical Center

Savannah Obregon, Vanderbilt University Medical Center

Dirk Orozco, Vanderbilt University Medical Center

Nelson Prato, Vanderbilt University Medical Center

Max Rohde, Vanderbilt University Medical Center

Russell Rothman, Vanderbilt University Medical Center

Jana Shirey-Rice, Vanderbilt University Medical Center

Krista Vermillion, Vanderbilt University Medical Center

Jacob Smith, Vanderbilt University Medical Center

Thomas Stewart, Vanderbilt University Medical Center / University of Virginia (Lead statistician)

Hsi-nien Tan, Vanderbilt University Medical Center

Meghan Vance, Vanderbilt University Medical Center

Maria Weir, Vanderbilt University Medical Center

**ACTIV-6 Site Investigators & Study Coordinators**

**Advanced Medical Care, Ltd:** Ray Bianchi, Jen Premas

**Ananda Medical Clinic:** Madhu Gupta, Greg Karawan, Santia Lima, Carey Ziomek

**Arena Medical Group:** Joseph Arena, Sonaly DeAlmeida

**Ascension St. John** Anuj Malik, Jane Bryce, Sarah Swint

**Chandler Regional Medical Center** Brian Tiffany, Charlotte Tanner, Allegra Sahelian

**Christ the King Health Care, P.C.:** Constance George-Adebayo, Adeolu Adebayo

**Christus St. Vincent Regional Medical Center:** Theresa Ronan, Ashley Woods, Christopher Gallegos, Tamara Flys, Olivia Sloan

**Clincept:** Anthony Olofintuyi, Joshua Samraj, Jackelyn Samraj, Amaya Averett

**Clinical Trials Center of Middle Tennessee:** Alex Slandzicki, Jessica Wallan

**Comprehensive Pain Management and Endocrinology:** Claudia Vogel, Sebastian Munoz

**David Kavtaradze MD, Inc.:** David Kavtaradze, Casandra Watson

**David Singleton MD, PA:** David Singleton, Marcus Sevier, Maria Rivon

**DHR Health Institute for Research:** Sohail Rao, Luis Cantu

**Diabetes and Endocrinology Assoc. of Stark County:** Arvind Krishna, Heidi Daugherty, Brandi Kerr, Kathy Evans

**Doctors Medical Group of Colorado Springs, P.C.:** Robert Spees, Mailyn Marta

**Duke University:** G. Michael Felker, Meaghan Beauchaine, Amanda Harrington, Daniah Amir, Juan Collazo

**Duke University Hospital:** Rowena Dolor, Lorraine Vergara, Jackie Jordan

**Elite Family Practice:** Valencia Burruss, Terri Hurst

**Emory University:** Paulina A. Rebolledo Esteinou, Igho Ofotokun, Cecilia Zhang, Jessica Traenkner, Mary M. Atha

**Essential Medical Care, Inc.:** Vickie James, Marcella Rogers

**Family Practice Doctors P.A.:** Chukwuemeka Oragwu, Ngozi Oguego

**First Care Medical Clinic:** Rajesh Pillai, Santia Lima

**Focus Clinical Research Solutions:** Ahab Gabriel, Emad Ghaly, Marian Michal

**G&S Medical Associates, LLC:** Gammal Hassanien, Samah Ismail, Yehia Samir

**George Washington University Hospital:** Andrew Meltzer, Ryan S. Heidish, Aditya Loganathan

**Geriatrics and Medical Associates:** Scott Brehaut, Angelina Roche

**GFC of Southeastern Michigan, PC:** Manisha Mehta, Nicole Koppinger

**Health Quality Primary Care:** Jose Baez, Ivone Pagan

**Highlands Medical Associates, P.A.:** Dallal Abdelsayed, Mina Aziz

**Hoag Memorial Hospital Presbyterian**: Philip Robinson, Grace Lozinski, Julie Nguyen

**HOPE Clinical Research:** Alvin Griffin, Michael Morris, Nicole Love, Bonnie Mattox, Raykel Martin

**Hugo Medical Clinic:** Victoria Pardue, Teddy Rowland

**Innovation Clinical Trials Inc.:** Juan Ruiz-Unger, Lionel Reyes, Navila Bacallao

**Jackson Memorial Hospital:** John Cienki

**Jadestone Clinical Research, LLC:** Jonathan Cohen, Ying Yuan, Jenny Li

**Jeremy W. Szeto, D.O., P.A.:** Jeremy Szeto

**Johns Hopkins University:** Mark Sulkowski, Lauren Stelmash, Sara Mekhael

**L&A Morales Healthcare, Inc:** Idania Garcia del Sol, Ledular Morales Castillo, Anya Gutierrez, Sabrina Prieto

**Lakeland Regional Medical Center:** Arch Amon, Andrew Barbera, Andrew Bugajski, Walter Wills, Kellcee Jacklin

**Lamb Health, LLC:** Deryl Lamb, Amron Harper

**Lapis Clinical Research:** Elmer Stout, Merischia Griffin

**Loyola University Medical Center:** Nina Clark, Mary Barsanti-Sekhar, Christina Carbrera-Mendez, Mary Rose Evans

**Maria Medical Center, PLLC:** Josette Maria, Oksana Raymond

**Medical Specialists of Knoxville:** Jeffrey Summers, Tammy Turner

**Medical University of South Carolina:** Leslie Lenert, Ebony Panaccione,

**Miller Family Practice, LLC:** Conrad Miller, Hawa Wiley

**Morehouse School of Medicine:** Austin Chan, Saadia Khizer

**North Shore University Health System/Evanston Hospital:** Nirav Shah, Oluwadamilola Adeyemi, Wei Ning Chi, July Chen, Melissa Morton-Jost

**Ochsner Clinic Foundation:** Julie Castex, Ali Quirch

**Olive View – UCLA Medical Center:** Hrishikesh Belani, Rosario Machicado, Bjorn Bjornsson

**Olivo Wellness Medical Center:** Jacqueline Olivo, Maria Maldonado

**Pine Ridge Family Medicine Inc.:** Anthony Vecchiarelli, Diana Gaytan-Alvarez

**Premier Health:** Vijaya Cherukuri, Santia Lima

**Providence Regional Medical Center:** Radica Alicic, Allison A. Lambert, Carissa Urbat, Joni Baxter, Ann Cooper

**Rapha Family Wellness:** Dawn Linn, Laura Fisher

**Raritan Bay Primary Care & Cardiology Associates:** Vijay Patel, Yuti Patel, Roshan Talati, Priti Patel

**Romancare Health Services:** Leonard Ellison, Angee Roman, Jeffrey Harrison

**Rush University Medical Center:** James Moy, Dina Naquiallah

**Spinal Pain and Medical Rehab, PC:** Binod Shah, Santia Lima

**Stanford University:** Prasanna Jagannathan, Upinder Singh, Orlando Quintero, Jake Scott, Andrew O’Donnell, Yasmin Jazayeri

**Sunshine Walk In Clinic:** Anita Gupta, N. Chandrasekar

**Superior Clinical Research:** Clifford Curtis, Briana White, Martha Dockery

**Tabitha B. Fortt, M.D., LLC:** Tabitha Fortt, Anisa Fortt

**Tampa General Hospital:** Jason Wilson, Jackie Marcelin, Brenda Farlow

**Temple University Hospital:** Nina Gentile, Casey Grady

**Texas Health Physicians Group:** Randall Richwine, Penny Pazier

**Texas Tech University Health Sciences Center in El Paso:** Edward Michelson, Susan Watts, Diluma Kariyawasam, Leann Rodriguez

**The Angel Medical Research:** Jose Luis Garcia, Ismarys Manresa, Angel A. Achong, Mari C. Garcia

**Trident Health Center:** Arvind Mahadevan, Santia Lima

**UF Health Precision Health Research:** Carla VandeWeerd, Jeffrey Lowenkron, Erica Sappington, Mitchell Roberts

**UMass Memorial Medical Center:** Jennifer Wang, Melissa Adams, Xinyi Ding, Mary Co

**University Diagnostics and Treatment Clinic:** Mark D'Andrea, Mina Aziz

**University Medical Center- New Orleans:** Stephen Lim, Madeline Young

**University of Arkansas:** Michael Wilson, Carly Eastin, Allyson Cheathem, Ahad Nadeem, Crystal Walters

**University of Cincinnati:** Margaret Powers-Fletcher, Douglas Brown, Delia Miller, Sylvere Mukunzi

**University of Florida-JAX-ASCENT:** Carmen Isache, Jennifer Bowman, Debra Martin

**University of Kansas – Wichita:** Brent Duran, Tiffany Schwasinger-Schmidt, Ashley Ast, Ashlie Cornejo, Allie Archer

**University of Miami:** Dushyantha Jayaweera, Maria Almanzar, Vanessa Motel

**University of Minnesota:** Matt Pullen, Neeta Bhat, Daniela Parra, Erica Urbina, Steven Arriaza

**University of Pittsburgh:** Akira Sekikawa, Emily Klawson, Jonathan Arnold, Nathan Weiland

**University of Texas Health Science Center at Houston:** Luis Ostrosky-Zeichner, Bela Patel, Virginia Umana, Laura Nielsen, Carolyn Z. Grimes

**University of Texas Health Science Center at San Antonio:** Thomas F. Patterson, Bridgette T. Soileau

**University of Virginia Health System:** Patrick E. H. Jackson, Heather M. Haughey

**Vaidya MD PLLC:** Bhavna Vaidya-Tank, Cameron Gould

**Vanderbilt University Medical Center:** Parul Goyal, Haley Pangburn, Lori Michalowski

**Wake Forest University Health Sciences:** John Williamson, Brittany Wortham, Rica Abbott,

**Weill Cornell Medical College:** Matthew McCarthy, Unwana Umana, Candace Alleyne, Britta Witting

**Well Pharma Medical Research:** Eddie Armas, Ramon O. Perez Landaburo, Michelle De La Cruz, Martha Ballmajo, Jorge Alvarez

**Supplemental Methods**

**Participant Monitoring**

The daily and follow-up assessments were monitored, and sites were actively notified of events requiring review, including serious adverse events (SAEs). In addition, participants were invited during assessments to request contact from the study team or to report any unusual circumstances. Failure to complete daily assessments also triggered a review for any possible SAEs. A missed assessment on the day after receiving the first dose of study medication (day 2) or any day of missed assessments up to day 14 prompted a notification to the site to contact the participant. All participants were instructed to self-report concerns either via an online event reporting system, by calling the site, or by calling a 24-hour hotline. Hospitalizations, a record of seeking other healthcare, or serious adverse events were extracted by site personnel from the participant’s medical record. Medical occurrences occurring before the receipt of study drug/placebo but after obtaining informed consent were not considered an adverse event.

#### **Independent Data Monitoring Committee Oversight**

The chair and/or convened committee reviewed data at least once monthly to review safety data, starting when the first participants met the 28-day timepoint:

| 10/19/2023 | full DSMB review |
| --- | --- |
| 10/31/2023 | Chair Review |
| 11/28/2023 | Chair Review |
| 12/21/2023 | Chair |
| 1/9/2024 | Chair Review |
| 1/29/2024 | Full |
| 2/20/2024 | Chair review |
| 3/5/2024 | Chair |
| 3/26/2024 | Full DSMB |
| 4/2/2024 | Chair |
| 4/24/2024 | Full DSMB |
| 5/14/2024 | Chair. |
| 6/11/2024 | Chair. |
| 7/9/2024 | Chair. |
| 7/16/2024 | Full DSMB: |
| 8/06/2024 | Chair review |
| 8/27/2024 | Chair |
| 9/19/2024 | Chair |
| 10/01/2024 | Chair review |
| 11/12/2024 | Chair review |
| 12/10/2024 | Chair review |

**Handling of Missing Data**

In both the primary and secondary endpoint analyses, missing data among covariates was addressed with conditional mean imputation. Approximately 3–9% of participants did not report activity level for the COVID clinical progression score endpoint at each time point, but the participants were known to be alive and at home. The missing activity levels was a type of interval censored outcome, as the participants were known to be either a 1 or 2 on the scale. The ordinal regression models were fit accounting for the interval censoring.

**Proportional Hazards**

The proportional hazards assumption of the primary endpoint was evaluated by generating visual diagnostics such as the log-log plot and plots of time-dependent regression coefficients for each predictor in the model, a diagnostic which indicates deviations from proportionality if the time-dependent coefficients are not constant in time.

**Heterogeneity of Treatment Effect Analysis**

For each characteristic, a proportional hazards regression model was constructed using the same covariates as the primary endpoint model plus additional interaction terms between treatment assignment and the characteristic of interest. To allow the possibility of non-linear trends along continuous characteristics, like age or calendar time, continuous covariates were included in the model as restricted cubic splines. The hazard ratios and 95% confidence intervals were calculated from asymptotic, model-based estimates at specific values. The continuous variables were not discretized into bins (or groups).

### **Overview of Agents and Enrollment Dates on the ACTIV-6 Platform**

1. Ivermectin 400 μg/kg daily for 3 days (June 23, 2021–February 4, 2022).^1^
2. Fluvoxamine 50 mg twice daily for 10 days (August 6, 2021–May 27, 2022).^2^
3. Inhaled fluticasone furoate 200 μg daily for 14 days (August 6, 2021–February 9, 2022).^3^
4. Ivermectin 600 μg/kg daily for 6 days (February 16, 2022–July 22, 2022).^4^
5. Fluvoxamine 50 mg twice daily on day 1 followed by 100 mg twice daily for 12 days (August 25, 2022–January 20, 2023).^5^
6. Montelukast 10 mg once daily for 14 days (January 27, 2023–June 23, 2023).^6^
7. Metformin (September 19, 2023–May 1, 2024).*

- 500 mg once per day for 1 day.
- Then 500 mg in the AM and 500 mg in the PM for 4 days.
- Then 500 mg in in the AM and 1000 mg in the PM for 9 days, 14 days total (36 tablets).
- Recommend taking at the end of a small snack or meal.

*The SARS-CoV-2 JN-1 variant dominated during most of the metformin enrollment period.^7^ The L455S mutation in the receptor binding domain, the characteristic feature of the SARS-CoV-2 JN-1 variant, enhanced its infectivity, stability, and immune evasion compared to prior variants.^8,9^ Overall, however, the JN-1 variants were associated with lower severity of symptoms and reduced risk of hospitalization.^9^ Compared to the JN-1 variants, the subsequent FLiRT variants, characterized by their R346T and F456L binding domain mutations, had improved immune evasion but reduced infectivity.^10,11^ Interestingly, the R346T mutation, which increases the affinity of the viral spike protein for binding to the human ACE2, thereby enhancing immune evasion, is similar to that found in earlier Omicron subvariants.^12,13^

### **COVID-19 Ordinal Outcome Scale**

The COVID-19 outcomes for this trial are based on the World Health Organization’s Ordinal Scale for Clinical Improvement and will be collected via the online system and from the medical record. The following outcomes will be assessed as part of the COVID Clinical Progression Scale:

0. No clinical or virological evidence of infection
1. No limitation of activities
2. Limitation of activities
3. Hospitalized, no oxygen therapy
4. Hospitalized, on oxygen by mask or nasal prongs
5. Hospitalized, on non-invasive ventilation or high-flow oxygen
6. Hospitalized, on intubation and mechanical ventilation
7. Hospitalized, on ventilation + additional organ support – pressors, renal replacement therapy, extracorporeal membrane oxygenation
8. Death

### **eTable 1. Day 1 symptom prevalence and severity***

| **Variable** | **Metformin**  **(n = 1443)** | **Placebo**  **(n = 1548)** | **Overall**  **(n = 2991)** |
| --- | --- | --- | --- |
| Overall Symptom burden on study day 1, No./total (%) | | | |
| None | 68/1322 (5.14) | 66/1388 (4.76) | 134/2710 (4.94) |
| Mild | 610/1322 (46.14) | 660/1388 (47.55) | 1270/2710 (46.86) |
| Moderate | 610/1322 (46.14) | 617/1388 (44.45) | 1227/2710 (45.28) |
| Severe | 34/1322 (2.57) | 45/1388 (3.24) | 79/2710 (2.92) |
| Individual Symptoms on study day 1: | | | |
| Fatigue, No./total (%) |  |  |  |
| None | 265/1255 (21.12) | 254/1321 (19.23) | 519/2576 (20.15) |
| Mild | 452/1255 (36.02) | 515/1321 (38.99) | 967/2576 (37.54) |
| Moderate | 449/1255 (35.78) | 458/1321 (34.67) | 907/2576 (35.21) |
| Severe | 89/1255 (7.09) | 94/1321 (7.12) | 183/2576 (7.10) |
| Dyspnea, No./total (%) |  |  |  |
| None | 726/1254 (57.89) | 805/1321 (60.94) | 1531/2575 (59.46) |
| Mild | 379/1254 (30.22) | 376/1321 (28.46) | 755/2575 (29.32) |
| Moderate | 130/1254 (10.37) | 124/1321 (9.39) | 254/2575 (9.86) |
| Severe | 19/1254 (1.52) | 16/1321 (1.21) | 35/2575 (1.36) |
| Fever, No./total (%) |  |  |  |
| None | 889/1254 (70.89) | 974/1321 (73.73) | 1863/2575 (72.35) |
| Mild | 264/1254 (21.05) | 264/1321 (19.98) | 528/2575 (20.50) |
| Moderate | 94/1254 (7.50) | 75/1321 (5.68) | 169/2575 (6.56) |
| Severe | 7/1254 (0.56) | 8/1321 (0.61) | 15/2575 (0.58) |
| Cough, No./total (%) |  |  |  |
| None | 217/1254 (17.30) | 227/1322 (17.17) | 444/2576 (17.24) |
| Mild | 549/1254 (43.78) | 598/1322 (45.23) | 1147/2576 (44.53) |
| Moderate | 421/1254 (33.57) | 442/1322 (33.43) | 863/2576 (33.50) |
| Severe | 67/1254 (5.34) | 55/1322 (4.16) | 122/2576 (4.74) |
| Nausea, No./total (%) |  |  |  |
| None | 957/1254 (76.32) | 1013/1320 (76.74) | 1970/2574 (76.53) |
| Mild | 217/1254 (17.30) | 218/1320 (16.52) | 435/2574 (16.90) |
| Moderate | 68/1254 (5.42) | 71/1320 (5.38) | 139/2574 (5.40) |
| Severe | 12/1254 (0.96) | 18/1320 (1.36) | 30/2574 (1.17) |
| Vomiting, No./total (%) |  |  |  |
| None | 1164/1254 (92.82) | 1207/1322 (91.30) | 2371/2576 (92.04) |
| Mild | 65/1254 (5.18) | 79/1322 (5.98) | 144/2576 (5.59) |
| Moderate | 21/1254 (1.67) | 33/1322 (2.50) | 54/2576 (2.10) |
| Severe | 4/1254 (0.32) | 3/1322 (0.23) | 7/2576 (0.27) |
| Diarrhea, No./total (%) |  |  |  |
| None | 921/1254 (73.44) | 1011/1322 (76.48) | 1932/2576 (75.00) |
| Mild | 257/1254 (20.49) | 219/1322 (16.57) | 476/2576 (18.48) |
| Moderate | 63/1254 (5.02) | 73/1322 (5.52) | 136/2576 (5.28) |
| Severe | 13/1254 (1.04) | 19/1322 (1.44) | 32/2576 (1.24) |
| Body aches, No./total (%) |  |  |  |
| None | 405/1254 (32.30) | 398/1322 (30.11) | 803/2576 (31.17) |
| Mild | 435/1254 (34.69) | 492/1322 (37.22) | 927/2576 (35.99) |
| Moderate | 345/1254 (27.51) | 366/1322 (27.69) | 711/2576 (27.60) |
| Severe | 69/1254 (5.50) | 66/1322 (4.99) | 135/2576 (5.24) |
| Sore throat, No./total (%) |  |  |  |
| None | 599/1253 (47.81) | 592/1322 (44.78) | 1191/2575 (46.25) |
| Mild | 402/1253 (32.08) | 431/1322 (32.60) | 833/2575 (32.35) |
| Moderate | 211/1253 (16.84) | 246/1322 (18.61) | 457/2575 (17.75) |
| Severe | 41/1253 (3.27) | 53/1322 (4.01) | 94/2575 (3.65) |
| Headache, No./total (%) |  |  |  |
| None | 543/1254 (43.30) | 562/1321 (42.54) | 1105/2575 (42.91) |
| Mild | 412/1254 (32.85) | 431/1321 (32.63) | 843/2575 (32.74) |
| Moderate | 236/1254 (18.82) | 249/1321 (18.85) | 485/2575 (18.83) |
| Severe | 63/1254 (5.02) | 79/1321 (5.98) | 142/2575 (5.51) |
| Chills, No./total (%) |  |  |  |
| None | 865/1254 (68.98) | 911/1320 (69.02) | 1776/2574 (69.00) |
| Mild | 249/1254 (19.86) | 258/1320 (19.55) | 507/2574 (19.70) |
| Moderate | 116/1254 (9.25) | 129/1320 (9.77) | 245/2574 (9.52) |
| Severe | 24/1254 (1.91) | 22/1320 (1.67) | 46/2574 (1.79) |
| Nasal symptoms, No./total (%) | | | |
| None | 451/1252 (36.02) | 460/1322 (34.80) | 911/2574 (35.39) |
| Mild | 439/1252 (35.06) | 469/1322 (35.48) | 908/2574 (35.28) |
| Moderate | 301/1252 (24.04) | 310/1322 (23.45) | 611/2574 (23.74) |
| Severe | 61/1252 (4.87) | 83/1322 (6.28) | 144/2574 (5.59) |
| New loss of sense of taste or smell, No./total (%) | | | |
| None | 802/1254 (63.96) | 839/1322 (63.46) | 1641/2576 (63.70) |
| Mild | 251/1254 (20.02) | 265/1322 (20.05) | 516/2576 (20.03) |
| Moderate | 127/1254 (10.13) | 141/1322 (10.67) | 268/2576 (10.40) |
| Severe | 74/1254 (5.90) | 77/1322 (5.82) | 151/2576 (5.86) |

*The most frequent was mild cough (44.53%), followed by mild fatigue (37.54%), and mild body aches (35.99%).

### **eTable 2. Serious adverse events**

| **Variable** | **Measure** | **Metformin** | **Placebo** | **Overall** |
| --- | --- | --- | --- | --- |
| Serious adverse events^a^ | **N** |  |  |  |
| Anemia |  | 1 | 0 | 1 |
| Atypical pneumonia |  | 1 | 0 | 1 |
| Encephalopathy acute |  | 1 | 0 | 1 |
| Exacerbation of asthma |  | 0 | 1 | 1 |
| Flu |  | 1 | 0 | 1 |
| Myocardial infarction |  | 1 | 0 | 1 |
| Pneumonia |  | 1 | 0 | 1 |
| Syncope |  | 1 | 0 | 1 |
| Transient ischemic attack |  | 0 | 1 | 1 |
| Urinary tract infection bacterial |  | 0 | 1 | 1 |
| Bronchitis acute viral |  | 1 | 0 | 1 |

**^a^**Using all available follow-up

### **eTable 3a. Time to recovery**

|  |  |  | **Metformin** | | | **Placebo** | | |
| --- | --- | --- | --- | --- | --- | --- | --- | --- |
| Day 14 | Group | Unadjusted HR (95% CI) | Events | n | % | Events | n | % |
| mITT | all | 1.00 (0.91, 1.09) | 968 | 1416 | 68.4 | 1046 | 1504 | 69.5 |
| Days from Symptom Onset | ≤3 | 1.08 (0.90, 1.30) | 214 | 301 | 71.1 | 229 | 330 | 69.4 |
|  | >3 | 0.98 (0.88, 1.08) | 754 | 1115 | 67.6 | 817 | 1174 | 69.6 |
| Previously Infected | No | 1.15 (1.01, 1.31)* | 436 | 604 | 72.2 | 463 | 681 | 68.0 |
|  | Yes | 0.90 (0.80, 1.02) | 492 | 730 | 67.4 | 540 | 746 | 72.4 |
|  | unknown | |  | 82 |  |  | 77 |  |
| Body Mass Index | <25 | 1.02 (0.85, 1.23) | 219 | 317 | 69.1 | 246 | 361 | 68.1 |
|  | 25-30 | 1.01 (0.88, 1.15) | 390 | 535 | 72.9 | 443 | 594 | 74.6 |
|  | ≥30 | 0.99 (0.85, 1.14) | 359 | 564 | 63.7 | 357 | 549 | 65.0 |
| Day 28 | Group | HR (95% CI) | Events | n | % | Events | n | % |
| mITT | all | 0.99 (0.92, 1.07) | 1213 | 1416 | 85.7 | 1303 | 1504 | 86.6 |
| Days from Symptom Onset | ≤3 | 1.09 (0.93, 1.29) | 264 | 301 | 87.7 | 284 | 330 | 86.1 |
|  | >3 | 0.97 (0.89, 1.06) | 949 | 1115 | 85.1 | 1019 | 1174 | 86.8 |
| Previously Infected | No | 1.07 (0.95, 1.21) | 516 | 604 | 85.4 | 585 | 681 | 85.9 |
|  | Yes | 0.95 (0.85, 1.05) | 638 | 730 | 87.4 | 660 | 746 | 88.5 |
|  | unknown | |  | 82 |  |  | 77 |  |
| Body Mass Index | <25 | 1.04 (0.89, 1.23) | 280 | 317 | 88.3 | 311 | 361 | 86.1 |
|  | 25-30 | 1.00 (0.88, 1.13) | 467 | 535 | 87.3 | 531 | 594 | 89.4 |
|  | ≥30 | 0.97 (0.86, 1.11) | 466 | 564 | 82.6 | 461 | 549 | 84.0 |

*p for interaction is <0.05. *Abbreviations:* HR=hazard ratio, and a HR > 1.0 is in the direction of benefit for metformin; mITT = modified intention to treat. The denominator is 2,920 because 71 participants did not provide any follow-up.

### **eTable 3b. Healthcare encounters by day 28***

|  | Group | **Metformin** | | | **Placebo** | | |
| --- | --- | --- | --- | --- | --- | --- | --- |
|  |  | Events | n | % | Events | n | % |
| mITT | all | 20 | 1416 | 1.41 | 24 | 1504 | 1.60 |
| Days from Symptom Onset | <=3 | 8 | 301 | 2.66 | 4 | 330 | 1.21 |
|  | >3 | 12 | 1115 | 1.08 | 20 | 1174 | 1.70 |
| Previously Infected | No | 6 | 604 | 0.99 | 9 | 681 | 1.32 |
|  | Yes | 12 | 730 | 1.64 | 14 | 746 | 1.88 |
|  | unknown |  | 82 |  |  | 77 |  |
| Body Mass Index | <25 | 3 | 317 | 0.95 | 5 | 361 | 1.39 |
|  | 25-30 | 4 | 535 | 0.75 | 7 | 594 | 1.18 |
|  | >=30 | 13 | 564 | 2.30 | 12 | 549 | 2.19 |
| *Defined as ED visit for Covid-19, hospitalization, or death. There were no deaths. Hospitalizations were rare; healthcare encounters before day 14 were infrequent. Previous infection was missing in 5.8% of metformin, 5.1% of placebo. *Abbreviation:* mITT = modified intention to treat.  The denominator is 2,920 because 71 participants did not provide any follow-up. | | | | | | | |

### **eFigure 1. Time from symptom onset to receipt of drug**

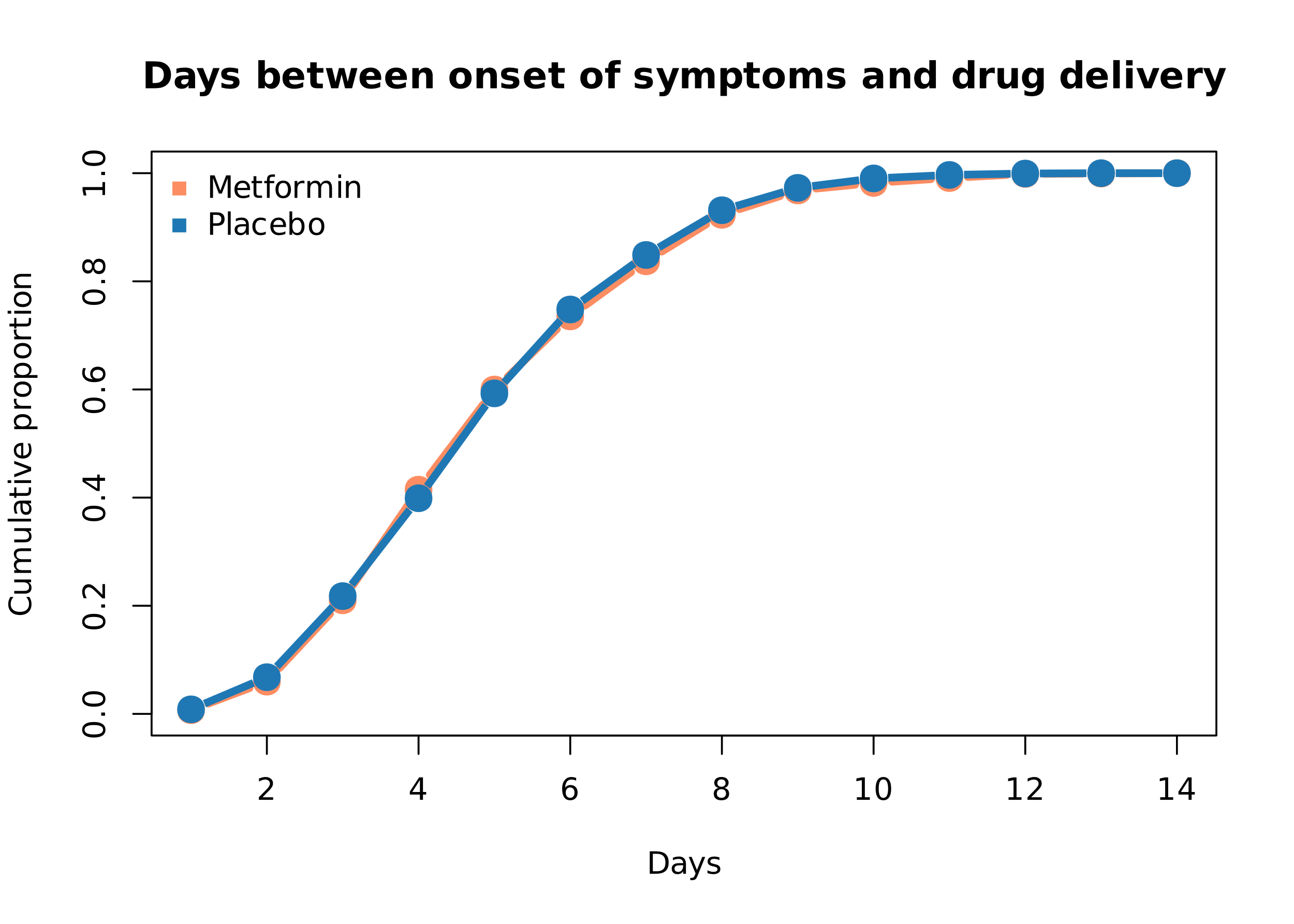

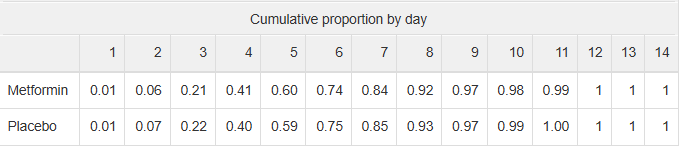

The time to study drug initiation from symptom onset is longer for those who did not start study drug on the day of delivery.

### **eFigure 2. Sensitivity analyses based on different patterns of missing symptom data**

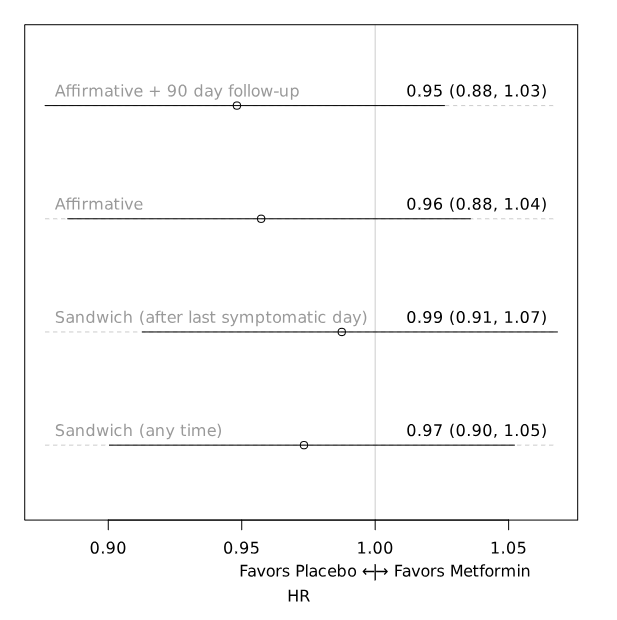

Sustained recovery is three consecutive symptom-free days. This ﬁgure illustrates the treatment eﬀect estimate for diﬀerent strategies of handling missing patient responses. "Aﬃrmative + 90 day follow-up" deﬁnition assumes that participants are symptomatic on the days without a response. The strategy does not censor recovery times earlier than day 28 if the day 90 survey is provided. "Aﬃrmative" assumes the same about days without a response. It diﬀers for the primary deﬁnition in that follow-up is only based on responses during the ﬁrst 28 days. A participant that fails to respond to surveys after day 14 is censored at day 14.

The second and third alternatives are imputation strategies, treating missing responses as symptom-free days if surrounded by symptom-free days. For instance, if a participant lacked a day two response but reported no symptoms on days one and three, the "Sandwich" deﬁnition marks the missing day as "no symptoms." The "Sandwich after last symptomatic day" deﬁnition only applies this rule after the last reported symptomatic day. The pattern of one or two days without symptoms followed by missing data was not assessed. The hazard ratios in the figure are from the covariate-adjusted, proportional hazards regression model without prior. Participants may have achieved 3 days without symptoms on or before day 1, and those individuals were not excluded.

### **eFigure 3. All-cause hospitalization or death; All-cause hospitalization or death, or clinic visit or emergency department visit**

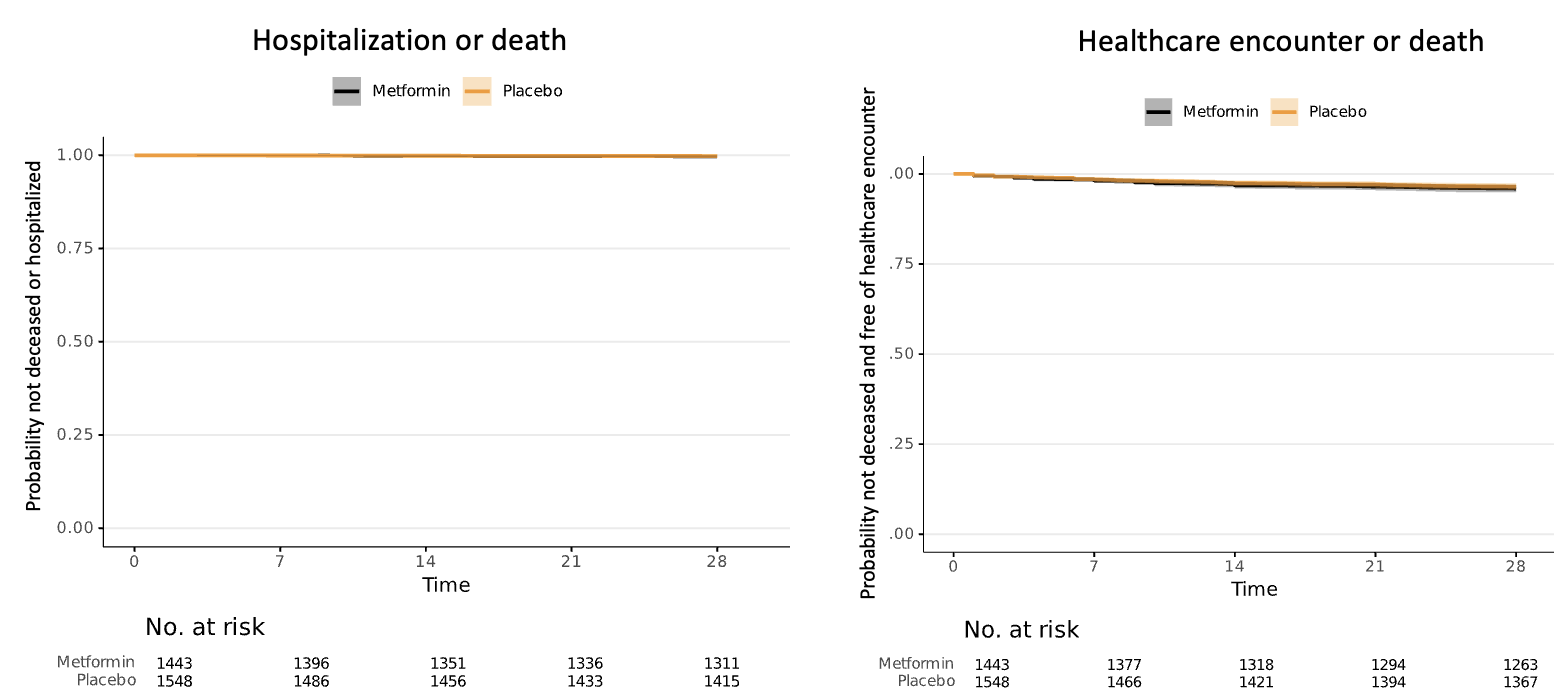

| Number Censored: | 128/1443 (8.9%) in metformin; 130/1548 (8.4%) in placebo. | 122/1443(8.5%) in metformin and 128/1548 (8.3%) in placebo |
| --- | --- | --- |

### **eFigure 4. Covid Clinical Progression Scale**

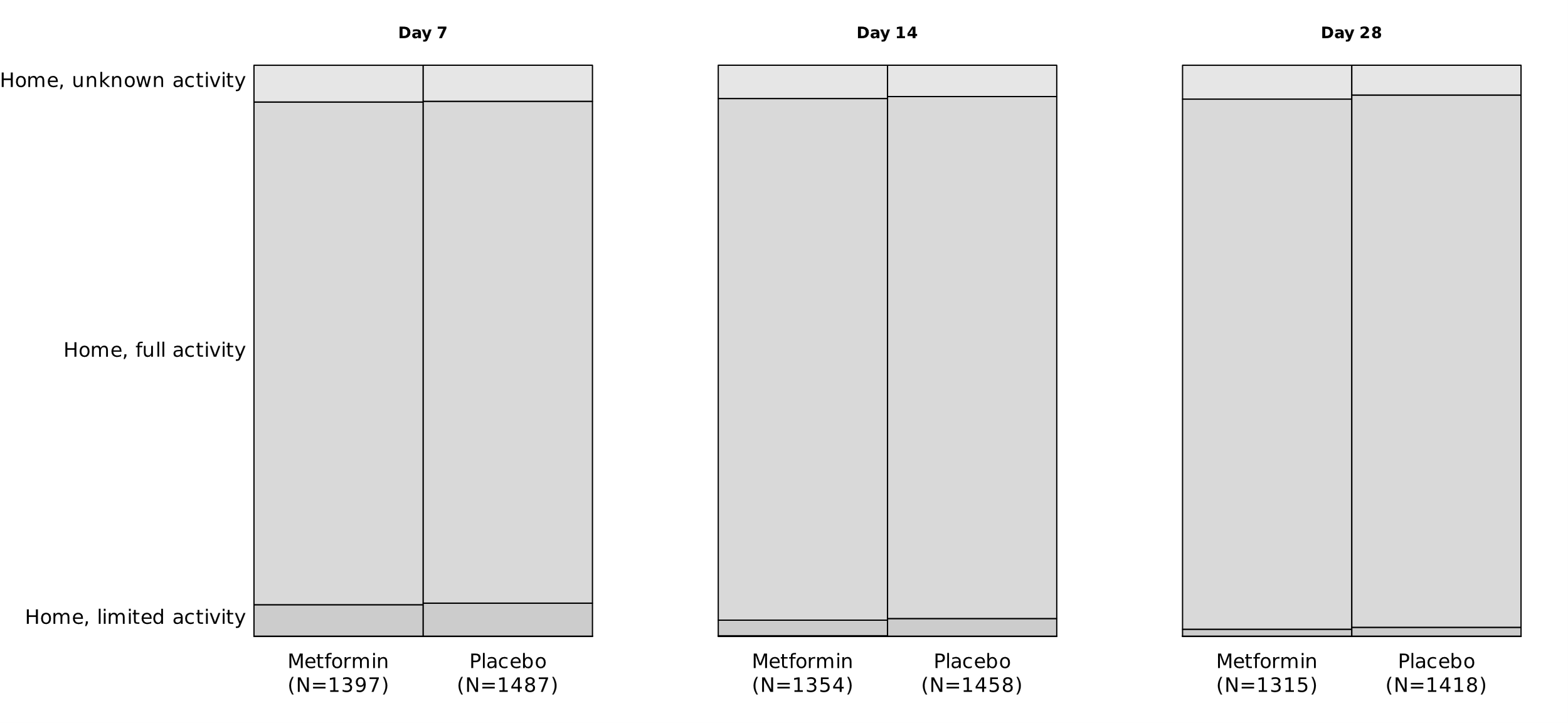

| Missing, n(%) | 46 (3.2) | 61 (3.9) | 89 (6.2) | 90 (5.8) | 128 (8.9) | 130 (8.4) |
| --- | --- | --- | --- | --- | --- | --- |

By Day 7, among responding participants, 2537/2700 (94%) participants reported Home, full activity and 163/2700 (6%) participants reported Home, limited activity.

### **eFigure 5. Heterogeneity of treatment effect for time to recovery**

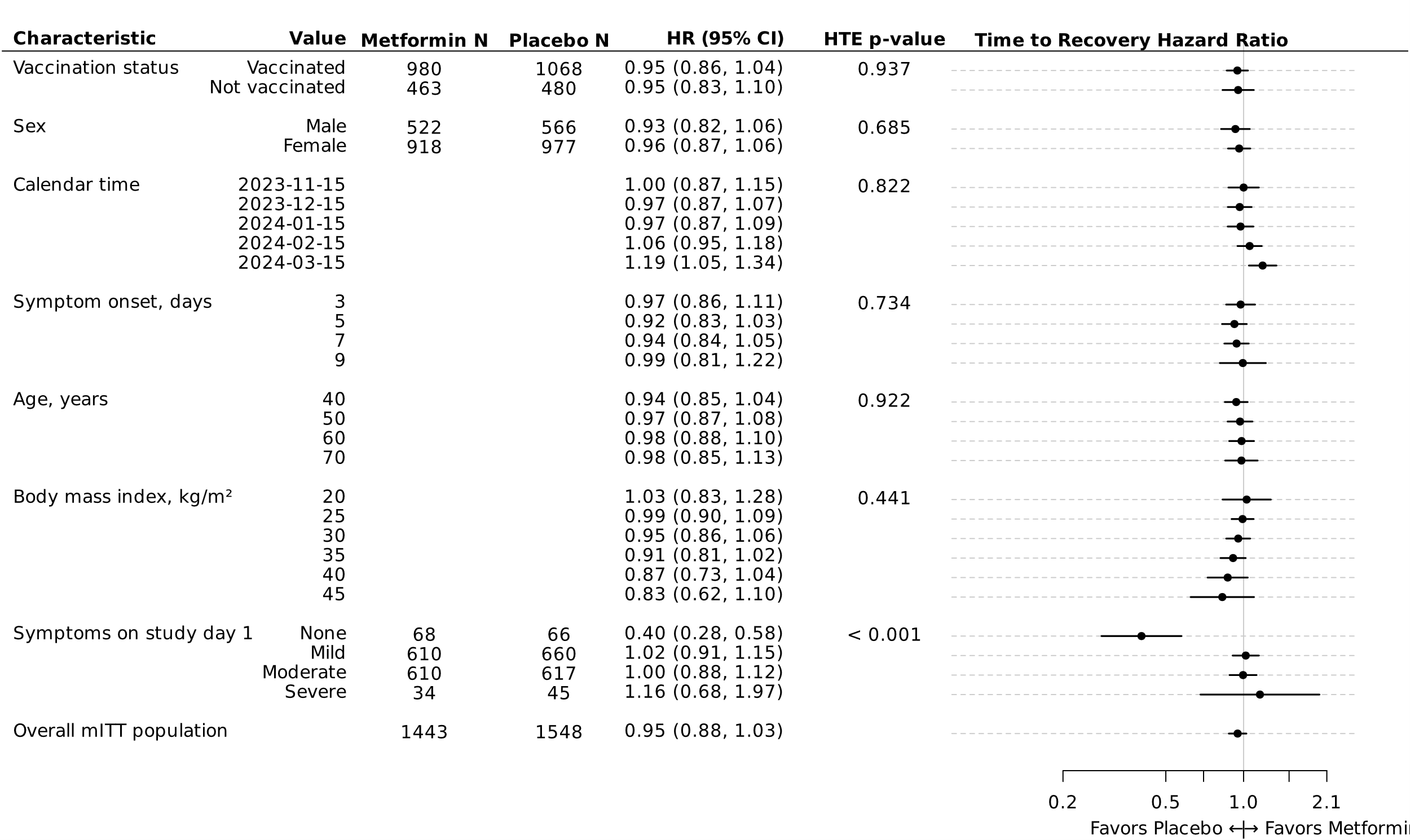

This ﬁgure presents the analyses exploring heterogeneity of treatment eﬀect derived from the primary endpoint—time to sustained recovery. These exploratory analyses, outlined in the statistical analysis plan, did not undergo multiplicity adjustment. A hazard ratio larger than 1 signiﬁes an accelerated recovery. Study day 1 corresponds to the participant's drug delivery day.

For each characteristic, a proportional hazards regression model was constructed using the same covariates as the primary endpoint model plus additional interaction terms between treatment assignment and the characteristic of interest. For example, the interaction of vaccination status and treatment assignment was added to the primary endpoint regression model to calculate a treatment eﬀect for the vaccinated and unvaccinated subgroups.

To allow the possibility of non-linear trends along continuous characteristics, like age or calendar time, the additional terms were interactions between treatment assignment and restricted cubic splines. Because the primary endpoint model did not include body mass index (BMI), the restricted cubic spline terms for BMI were also added to the model (sometimes call main eﬀects) in addition to the interaction terms. Because the primary endpoint model only included a single linear term for symptom onset, the nonlinear terms of the restricted cubic spline were also added to the model in addition to the interaction terms. The hazard ratios and 95% conﬁdence intervals were calculated from asymptotic, model-based contrasts. The hazard ratio for the full study population was generated from the primary endpoint model without prior adjustments. The estimates shown in the HTE plot are estimates calculated from the smooth, modeled relationship. Of the 2,991 in the mITT, 2,641 (88.3%) were enrolled as of March 15, 2024. March 2024 was the approximate date the FLiRT variants became dominant and aligns with the most chronologically recent category in the interaction term between treatment assignment and calendar time.

### **eFigure 6. Kaplan-Meier curves of sustained recovery by onset of symptoms and symptom severity on the day of delivery**

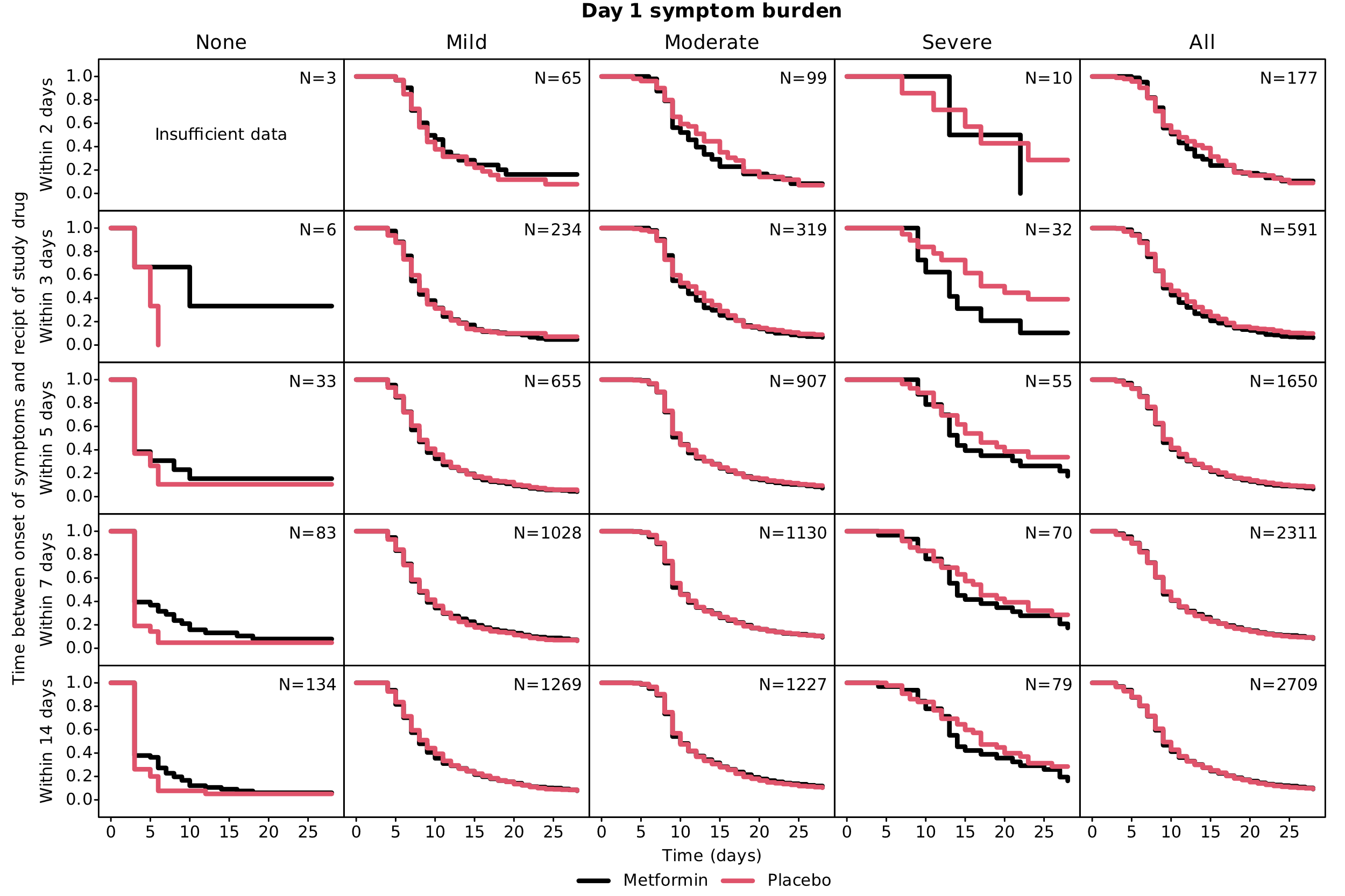

Of the 246 with severe symptoms on day 1, 17.1% received study drug within 3 days. Of the 259 with no symptoms on day 1, 3.5% received study drug within 3 days. Participants who achieved 3 days without symptoms on or before day 1 were not excluded.

### **eFigure 7. Kaplan-Meier curves of time to recovery stratified by symptom onset**

| 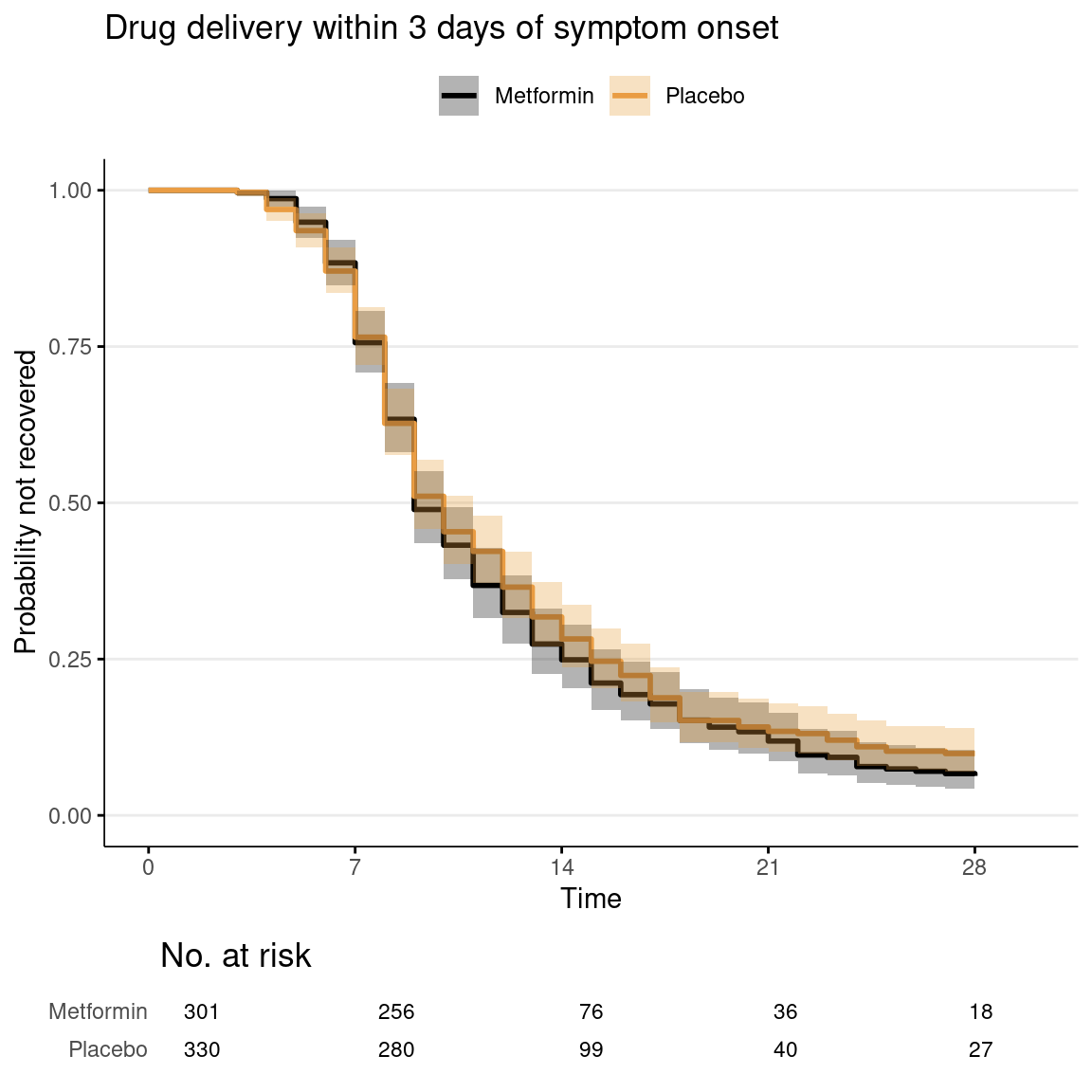 | 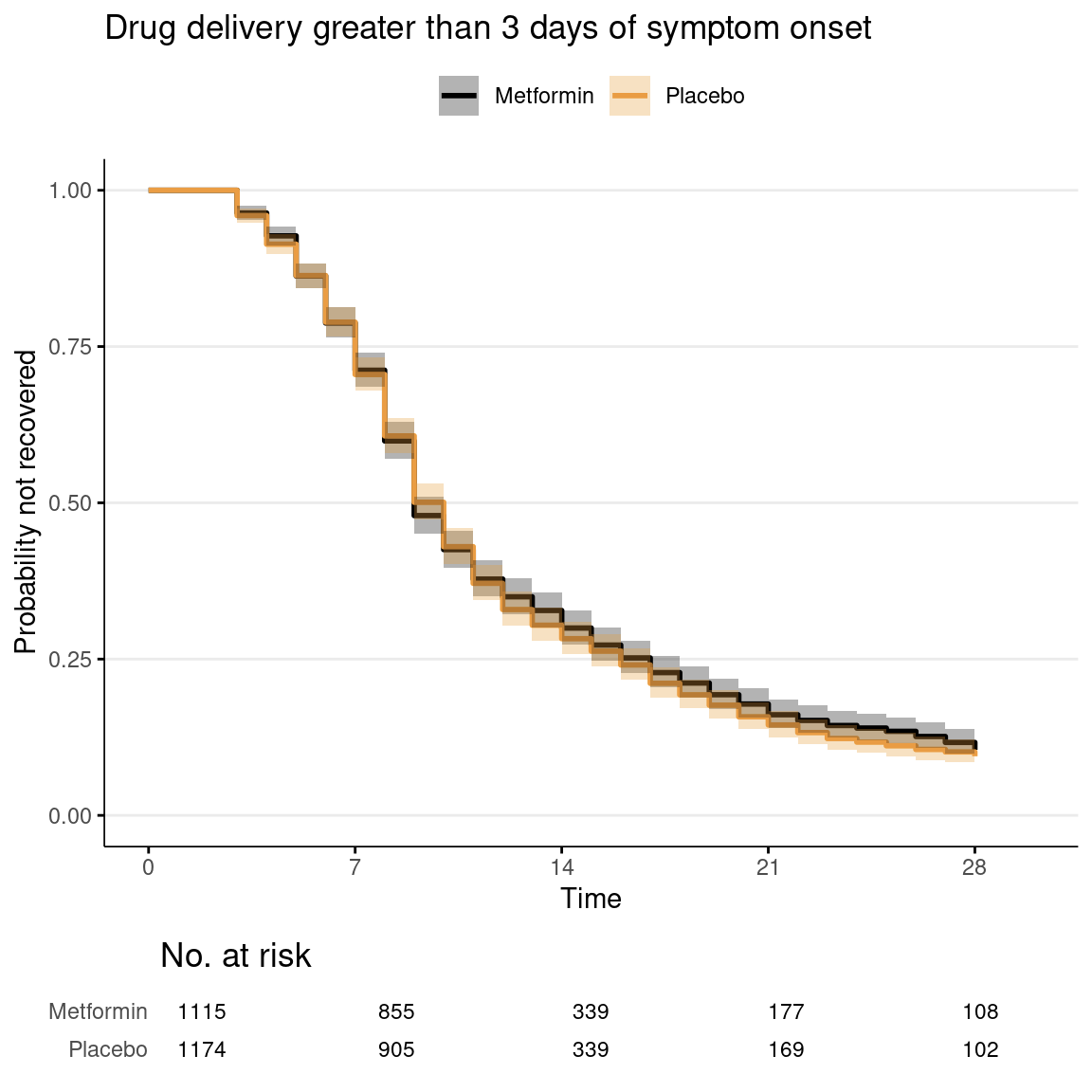 |
| --- | --- |

For those receiving study drug within 3 days of symptom onset, the median time to sustained recovery was 9 (95% CI 9 -10) in the metformin group and 10 (95% CI 9-11) in the placebo group.

**eFigure 8. Posterior density plots of all covariates in the primary outcome model**
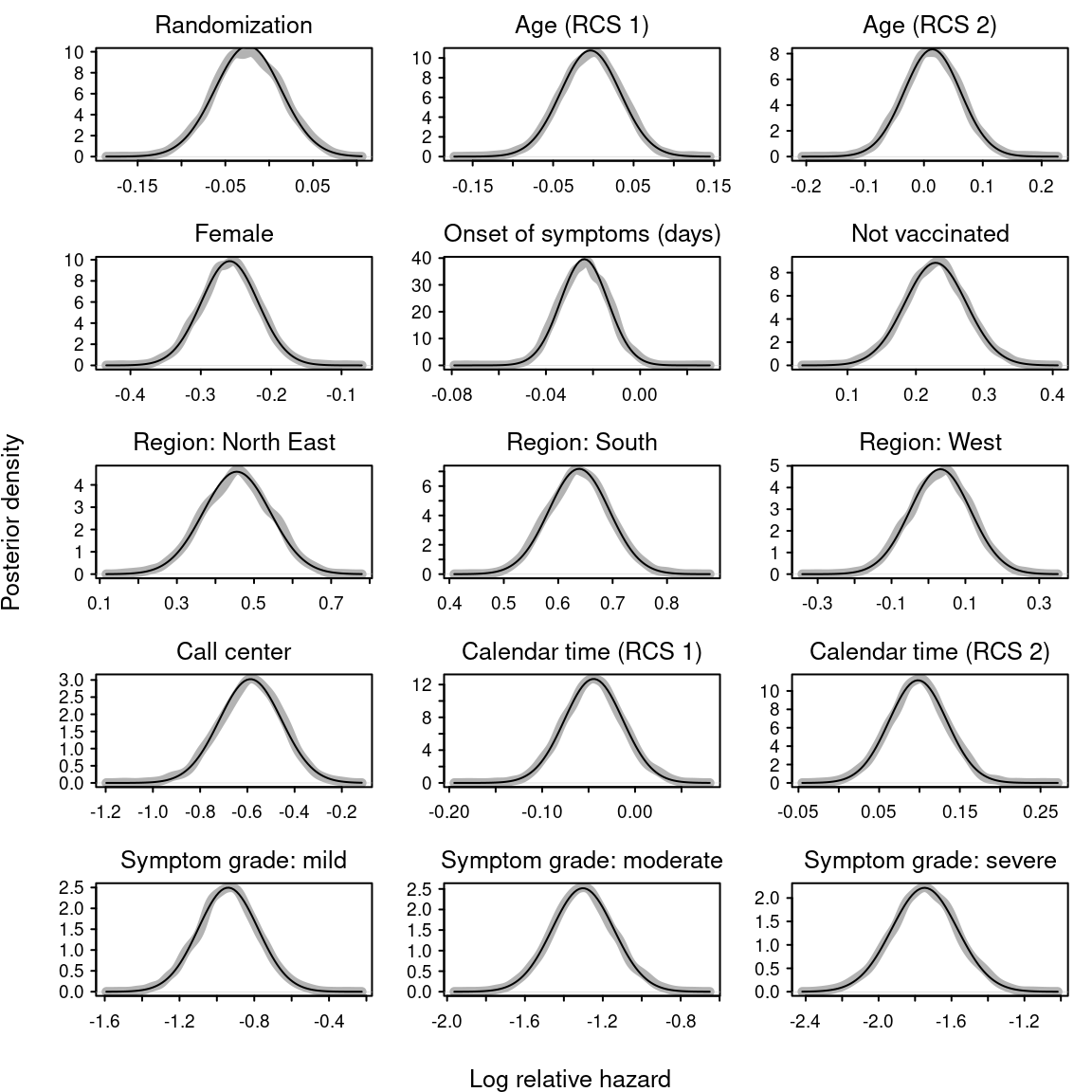

This ﬁgure provides the posterior densities of each regression coeﬃcient in the primary endpoint model. The x-axis is the log relative hazard; the x-axis is density. The thick, grey lines are the kernel density estimates; thin, black lines represent parametric normal density estimates of the same.
